# COVID-19 Associated Pulmonary Aspergillosis: Systematic Review and Patient-Level Meta-analysis

**DOI:** 10.1101/2021.05.21.21257626

**Authors:** Ruwandi M. Kariyawasam, Tanis C. Dingle, Brittany E. Kula, Wendy I. Sligl, Ilan S. Schwartz

## Abstract

**Rationale:** Pulmonary aspergillosis may complicate COVID-19 and contribute to excess mortality in intensive care unit (ICU) patients. The incidence is unclear because of discordant definitions across studies.

**Objective:** We sought to review the incidence, diagnosis, treatment, and outcomes of COVID-19-associated pulmonary aspergillosis (CAPA), and compare research definitions.

**Methods:** We systematically reviewed the literature for ICU cohort studies and case series including ≥ patients with CAPA. We calculated pooled incidence. Patients with sufficient clinical details were reclassified according to 4 standardized definitions (Verweij, White, Koehler, and Bassetti).

**Measurements:** Correlations between definitions were assessed with Spearman’s rank test. Associations between antifungals and outcome were assessed with Fisher’s Exact test.

**Main Results:** 38 studies (35 cohort studies and 3 case series) were included. Among 3,297 COVID-19 patients in ICU cohort studies, 313 were diagnosed with CAPA (pooled incidence 9.5%). 197 patients had patient-level data allowing reclassification. Definitions had limited correlation with one another (ρ=0.330 to 0.621, p<0.001). 38.6% of patients reported to have CAPA did not fulfil any research definitions. Patients were diagnosed after a median of 9 days (interquartile range 5-14) in ICUs. Tracheobronchitis occured in 5.3% of patients examined with bronchoscopy. The mortality rate (50.0%) was high, irrespective of antifungal use (p=0.28); this remained true even when the analysis was restricted to patients meeting standardized definitions for CAPA.

**Conclusions:** The reported incidence of CAPA is exaggerated by use of non-standard definitions. Further research should focus on identifying patients likely to benefit from antifungals.

## Introduction

Given the high morbidity and mortality associated with severe COVID-19 (1), it is imperative to rule out co-infections that may contribute to poor outcomes. In patients with severe COVID-19 requiring admission to the intensive care unit (ICU), pulmonary aspergillosis has been reported to be a relatively common and important complication contributing to increased mortality (2).

An association between viral respiratory tract infection and aspergillosis has been established in influenza: the incidence of influenza-associated pulmonary aspergillosis (IAPA) ranges from 7-30% within 3 days of ICU admission, and is associated with ∼50% mortality (3, 4). Similarly, aspergillosis has been reported to occur in up to a third of patients with critical COVID-19 (5–7). However, diagnosis and classification remains challenging.

Most reported cases of COVID-19 associated pulmonary aspergillosis (CAPA) have occurred in immunocompetent patients, and thus patients without histologic evidence of invasive fungal disease cannot meet the classic research definitions of the European Organization for Research and Treatment of Cancer - Mycoses Study Group Education and Research Consortium (EORTC-MSGERC) for probable aspergillosis, since these require the presence of immunocompromising host factors (8). Alternative research definitions for CAPA have been proposed, first modified from IAPA and subsequently refined given the rapid pace of data generation and knowledge synthesis (6,9–12).

A common limitation among these definitions is the characterization of CAPA by non-specific clinical and radiographic findings that are difficult to distinguish from COVID-19 alone. Moreover, mycological findings (such as detection of *Aspergillus* in respiratory samples by culture, antigen detection, or PCR) cannot reliably distinguish between infection and colonization.

The lack of a validated research case definition may result in missed or mis-identified individual cases, thereby delaying appropriate antifungal therapy or resulting in unnecessary therapy, both of which may result in poor outcomes. We conducted a systematic review of case series and cohort studies assessing patients diagnosed with CAPA while in the ICU and report incidence, diagnostics, treatments and mortality.

## Methods

### Protocol and Registration

The review protocol has been registered in PROSPERO International Prospective Register of Systematic Reviews, CRD42020204123. The review and reporting were conducted according to PRISMA guidelines.

### Eligibility Criteria

We included cohort studies of patients with COVID-19 admitted to ICUs who were evaluated for pulmonary aspergillosis using fungal diagnostics (including direct microscopy, fungal culture, *Aspergillus* PCR, and/or galactomannan testing on respiratory tract specimens, and/or galactomannan or 1,3-β-D-glucan (BDG) testing in blood), and case series of ICU patients with CAPA. We excluded case reports with fewer than 3 patients.

### Information Sources

We searched 3 electronic databases (PubMed, Embase and Web of Science) from database inception to March 13, 2021 irrespective of language to identify relevant studies. A separate search was conducted on medRxiv to search for studies posted ahead of peer-review. The search strategy was restricted to humans. Reference lists and cited bibliographies were also hand searched. Two reviewers assessed for study eligibility, selected studies and extracted data. Where assessments were discordant and could not be settled by discussion between the reviewers, these were arbitrated by a third reviewer.

### Search

The following search strategy was employed on PubMed from inception to March 13, 2021: ((“aspergillus”[All Fields]) OR (“aspergillosis”[All Fields]))) AND ((((“sars cov2”[All Fields]) OR (“covid”[All Fields])) OR (“2019 ncov”[All Fields])) OR (“novel coronavirus”[All Fields]). Similar search terms were used for the other databases.

### Definitions

Several research definitions for CAPA have been proposed (Table 1). We analyzed these by extracting patient-level data and reclassifying patients according to 4 research definitions: an expert consensus definition for influenza-associated pulmonary aspergillosis (IAPA) adapted to COVID-19 (Verweij) (10), the CAPA definitions from Wales (White) (2), expert consensus definitions of the European Confederation of Medical Mycology (ECMM) and the International Society for Human and Animal Mycoses (ISHAM) for CAPA (Koehler) (11), and the EORTC-MSGERC Intensive Care Working Group definitions for invasive aspergillosis in ICU patients (Bassetti) (12).

**Table 1.**
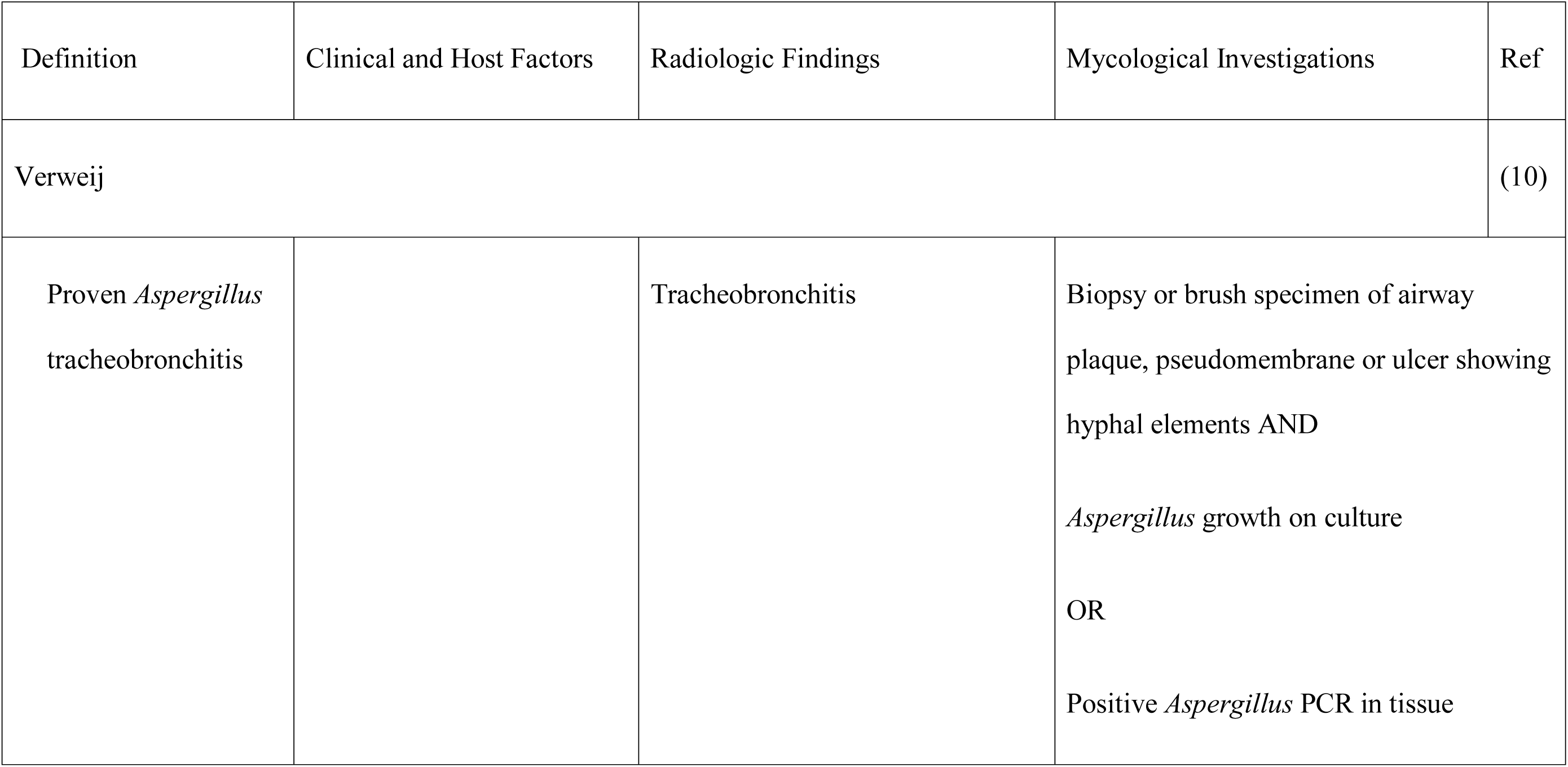
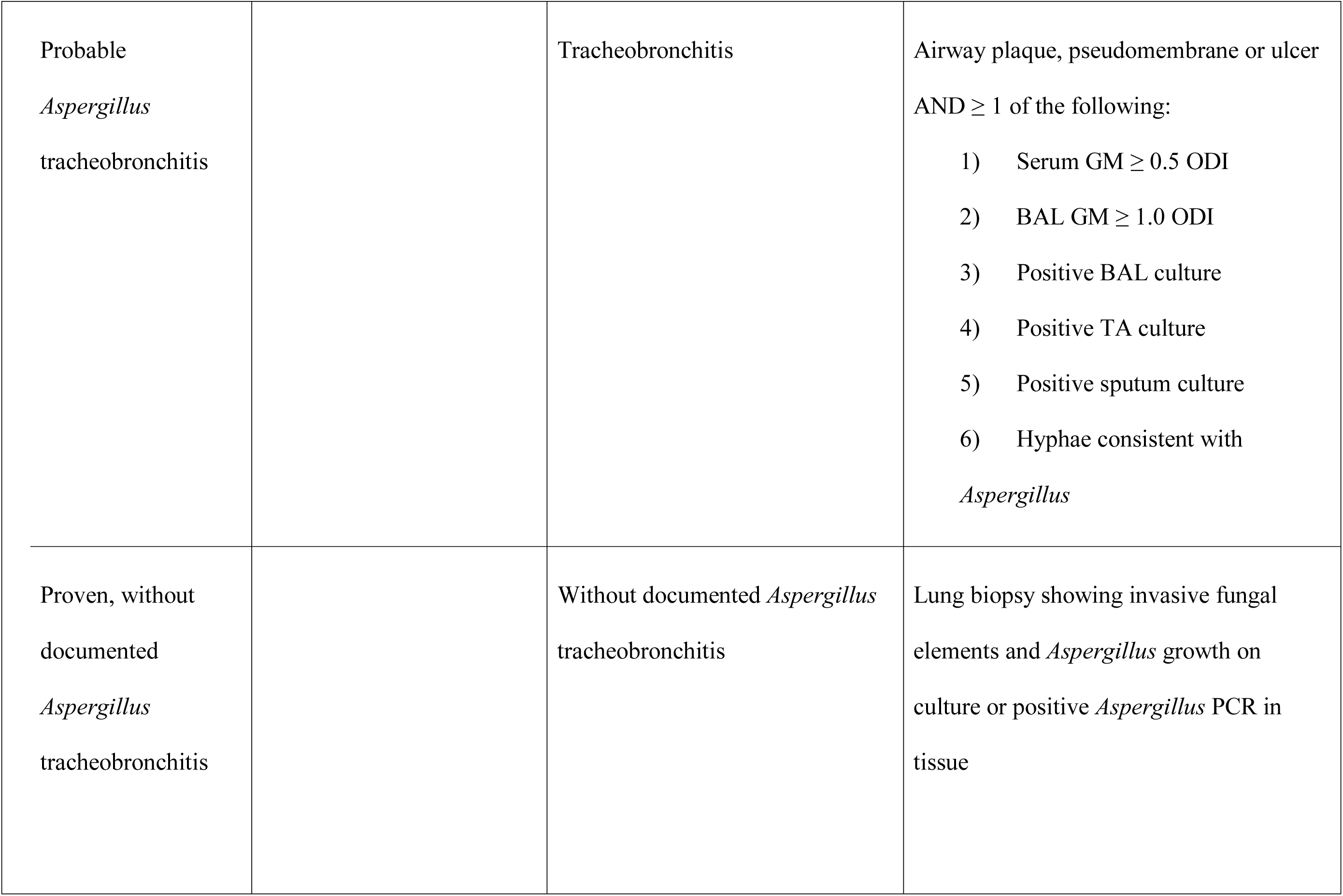
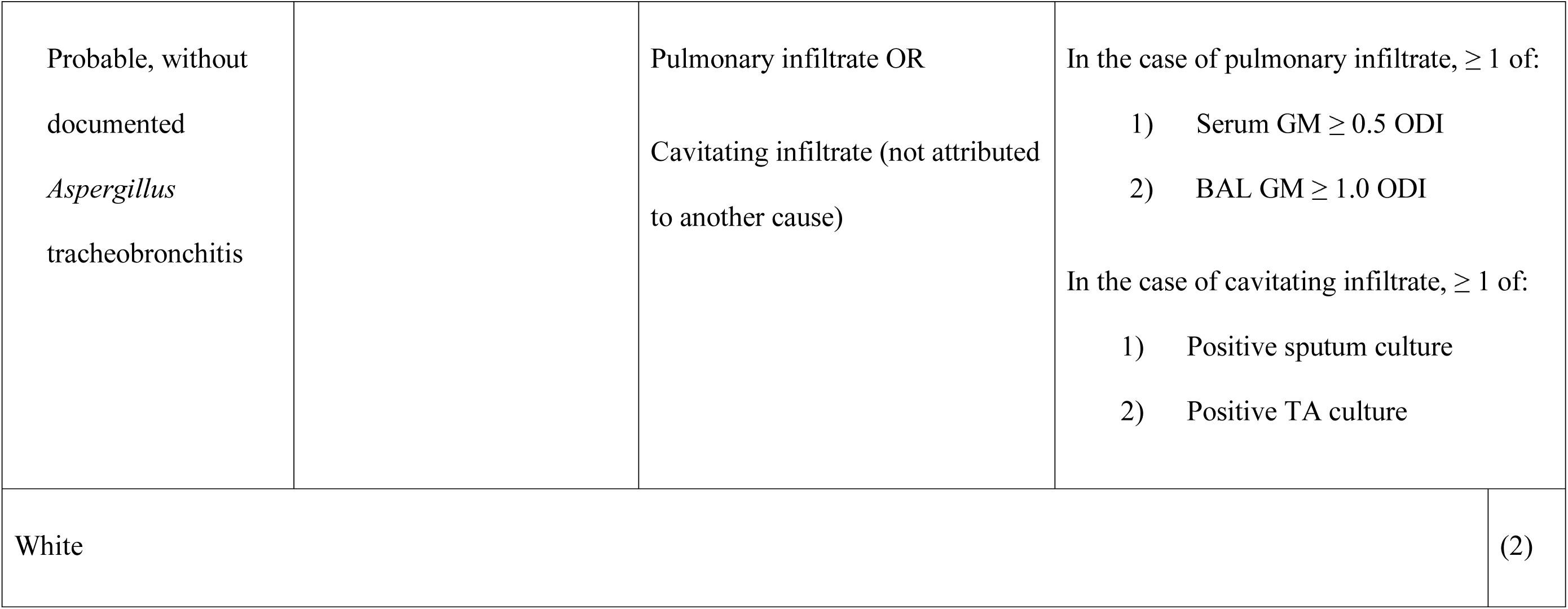
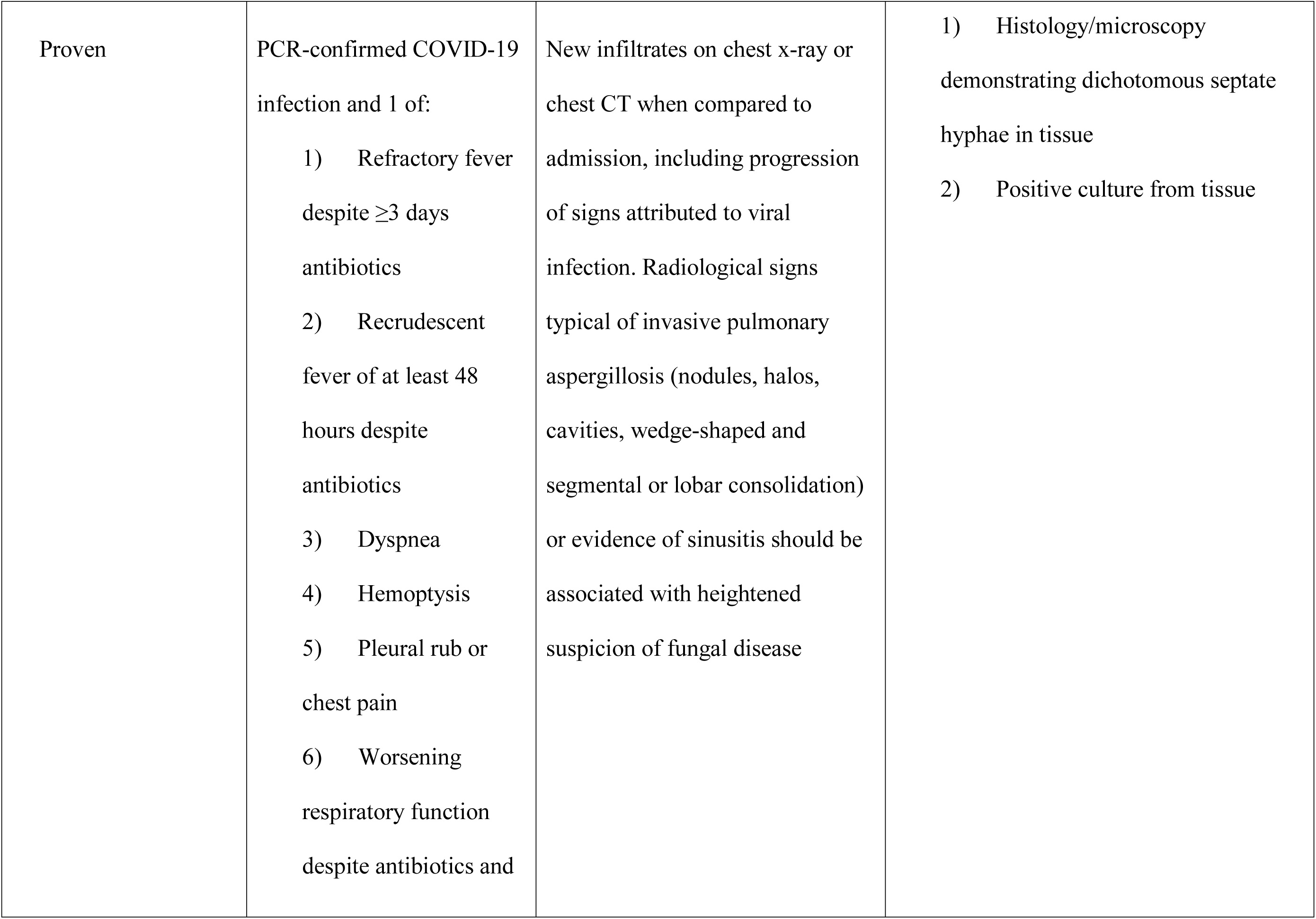
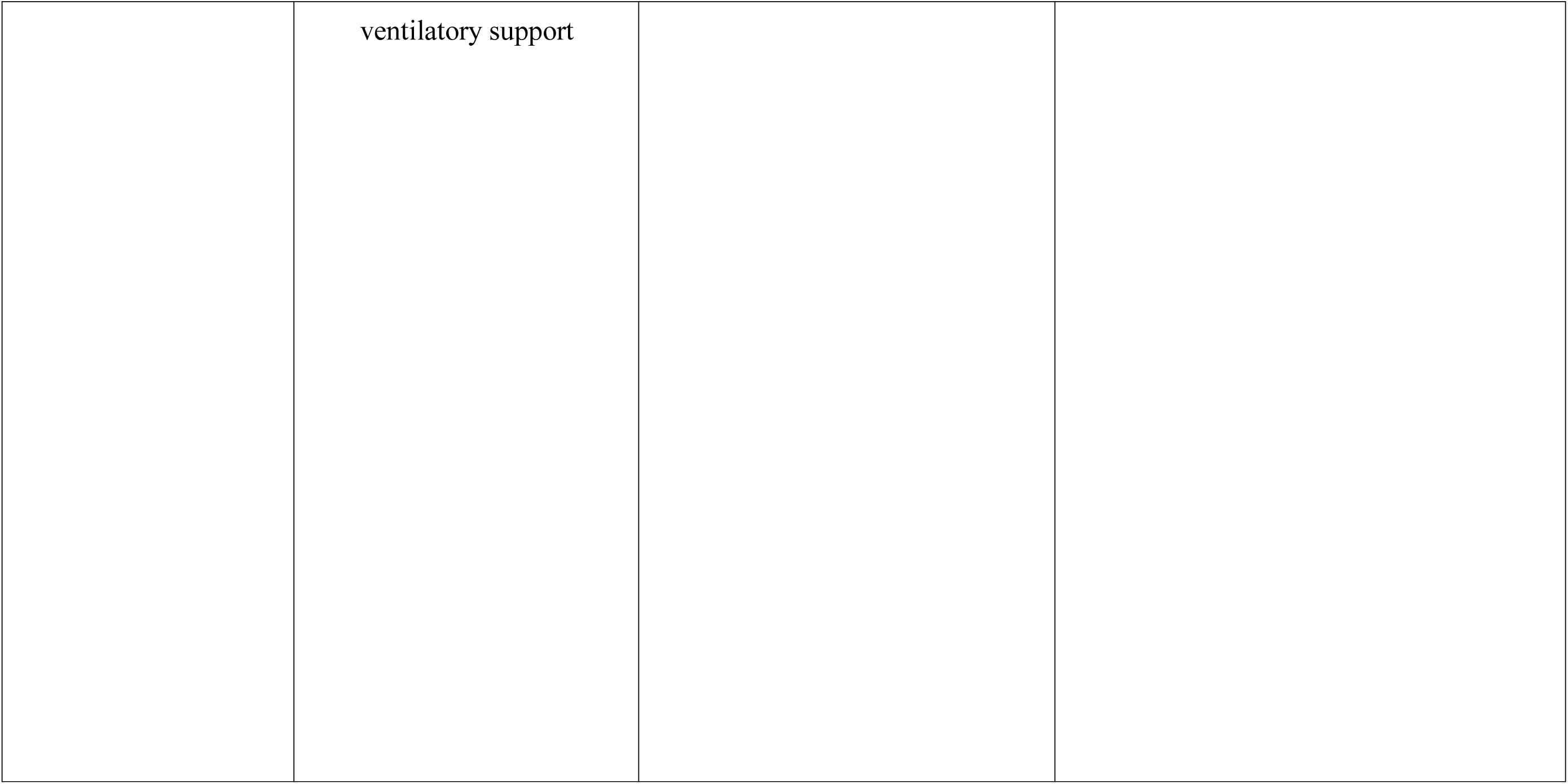
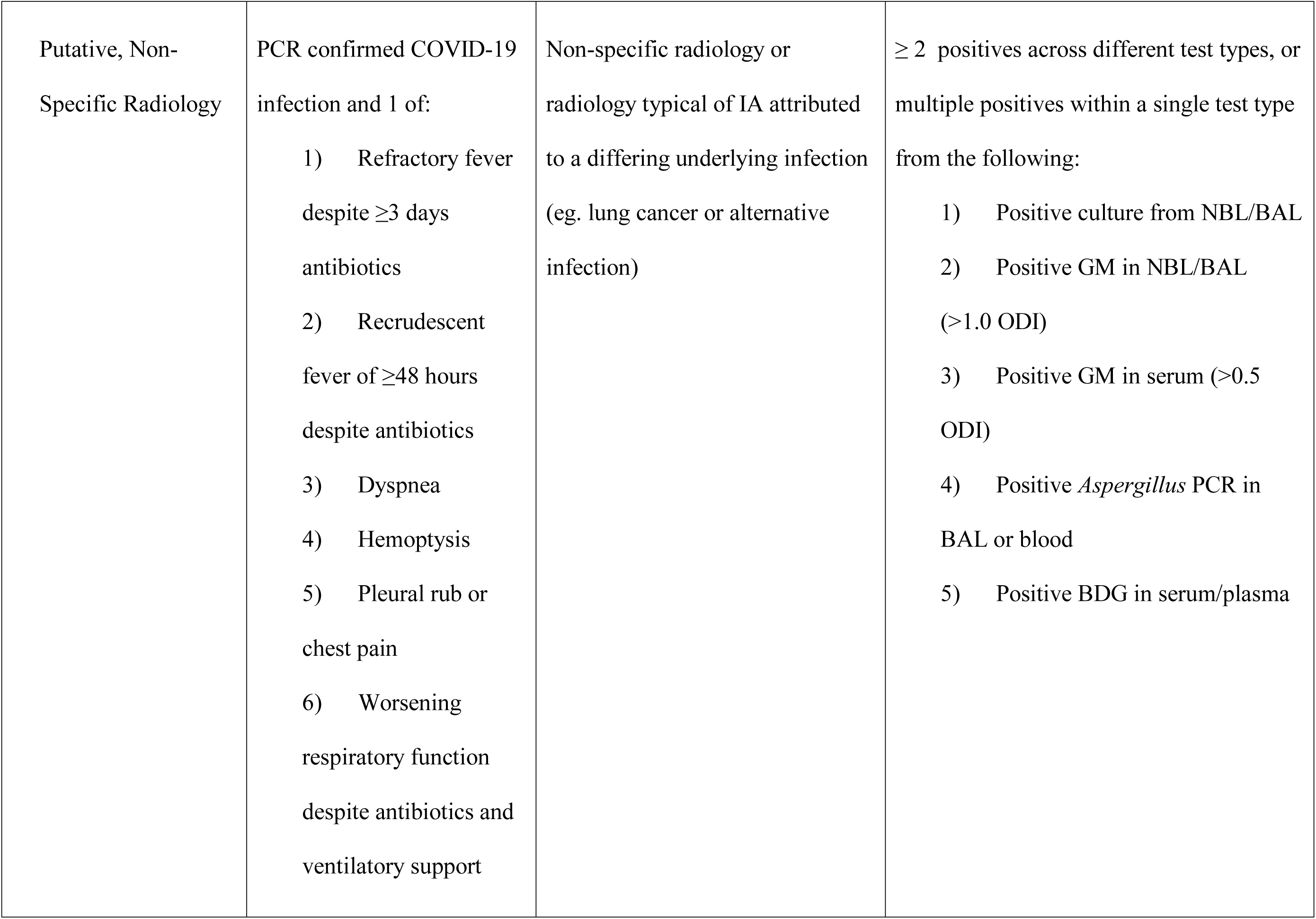
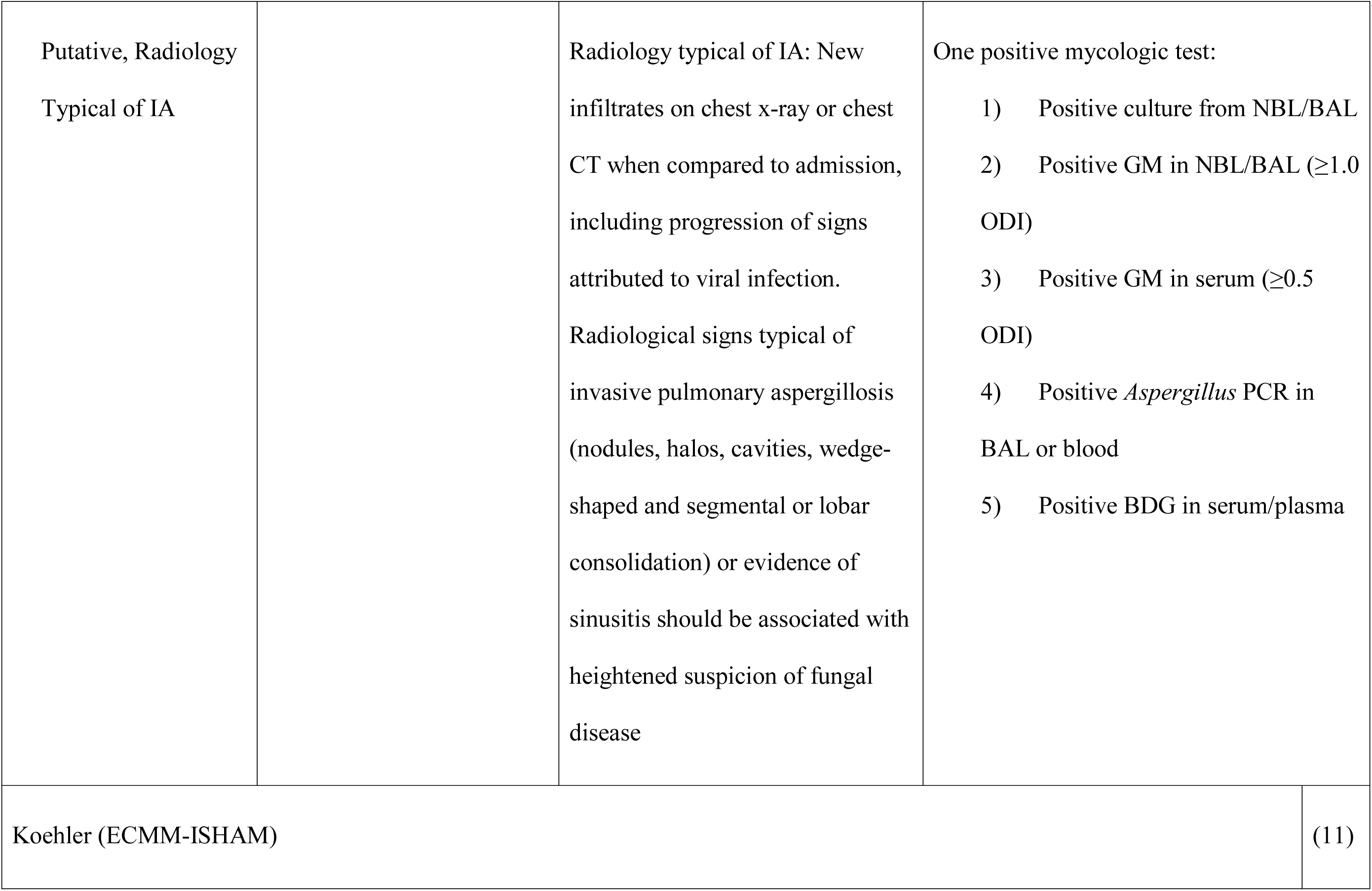
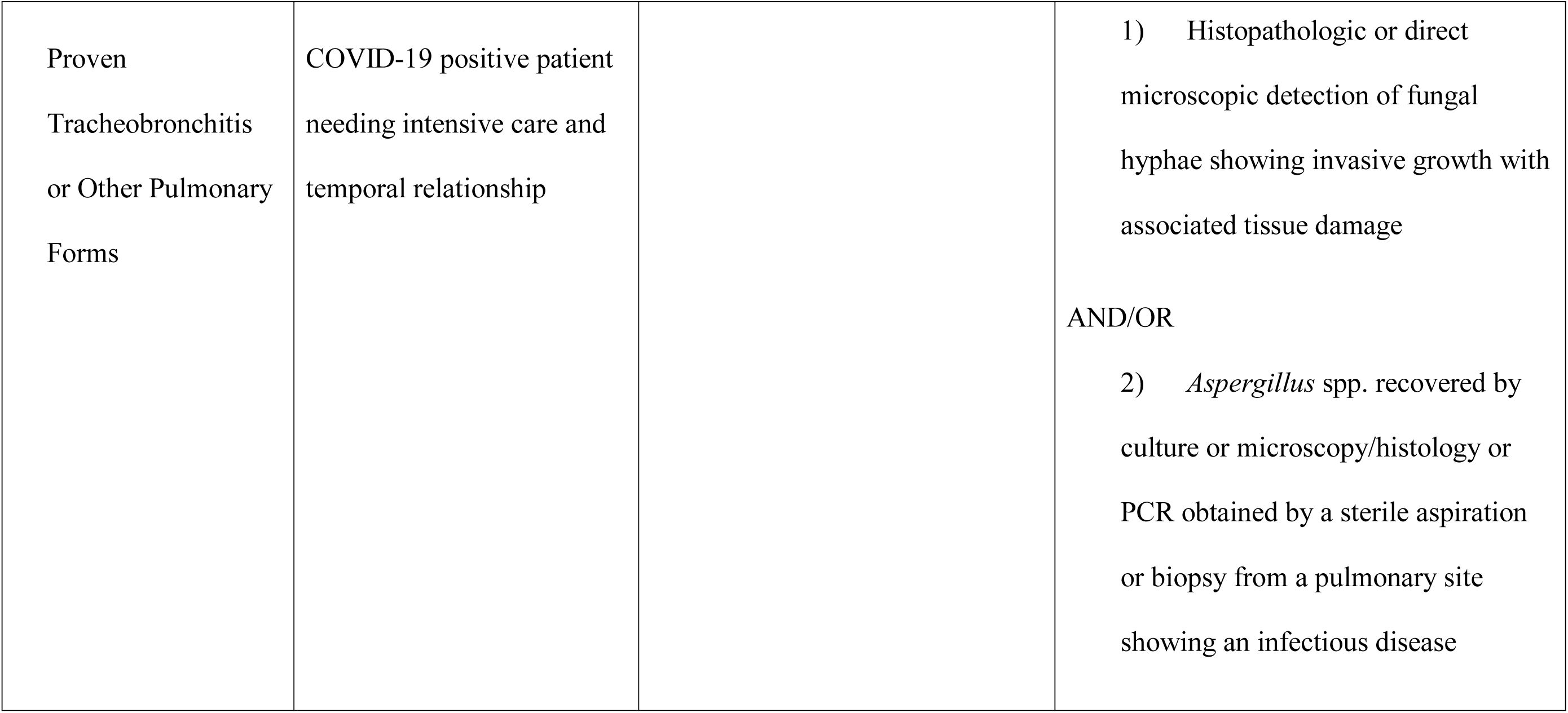
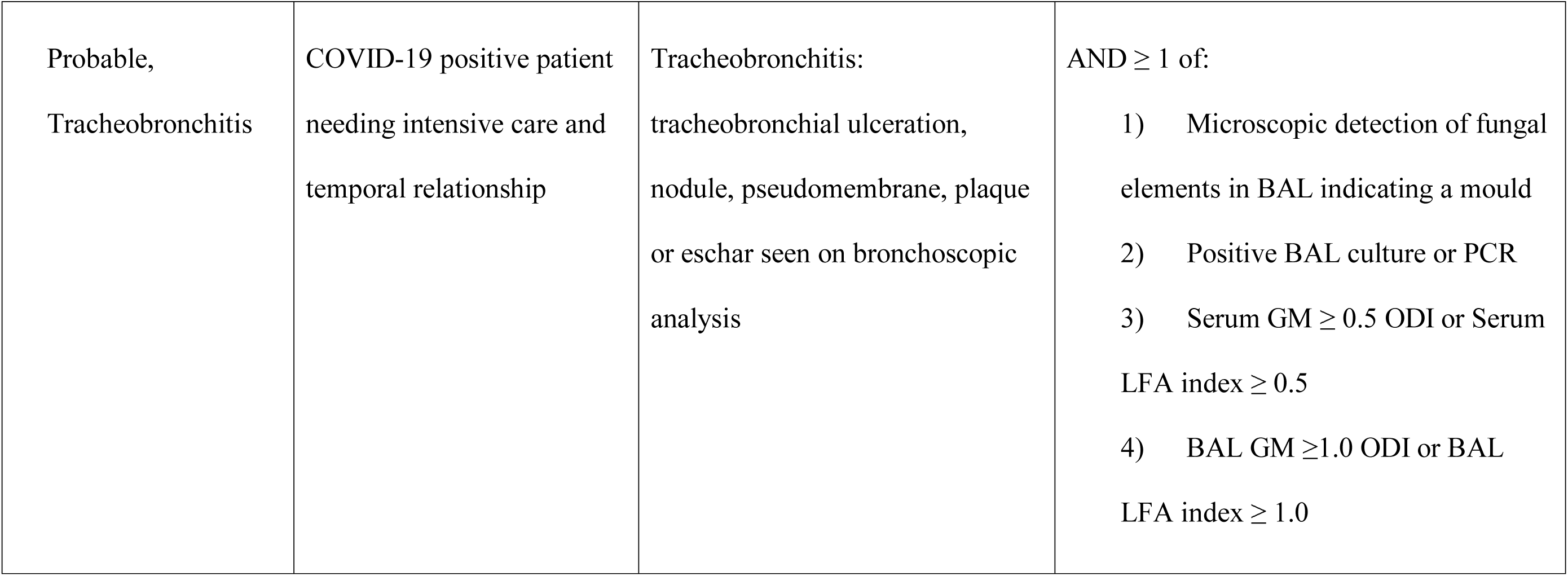
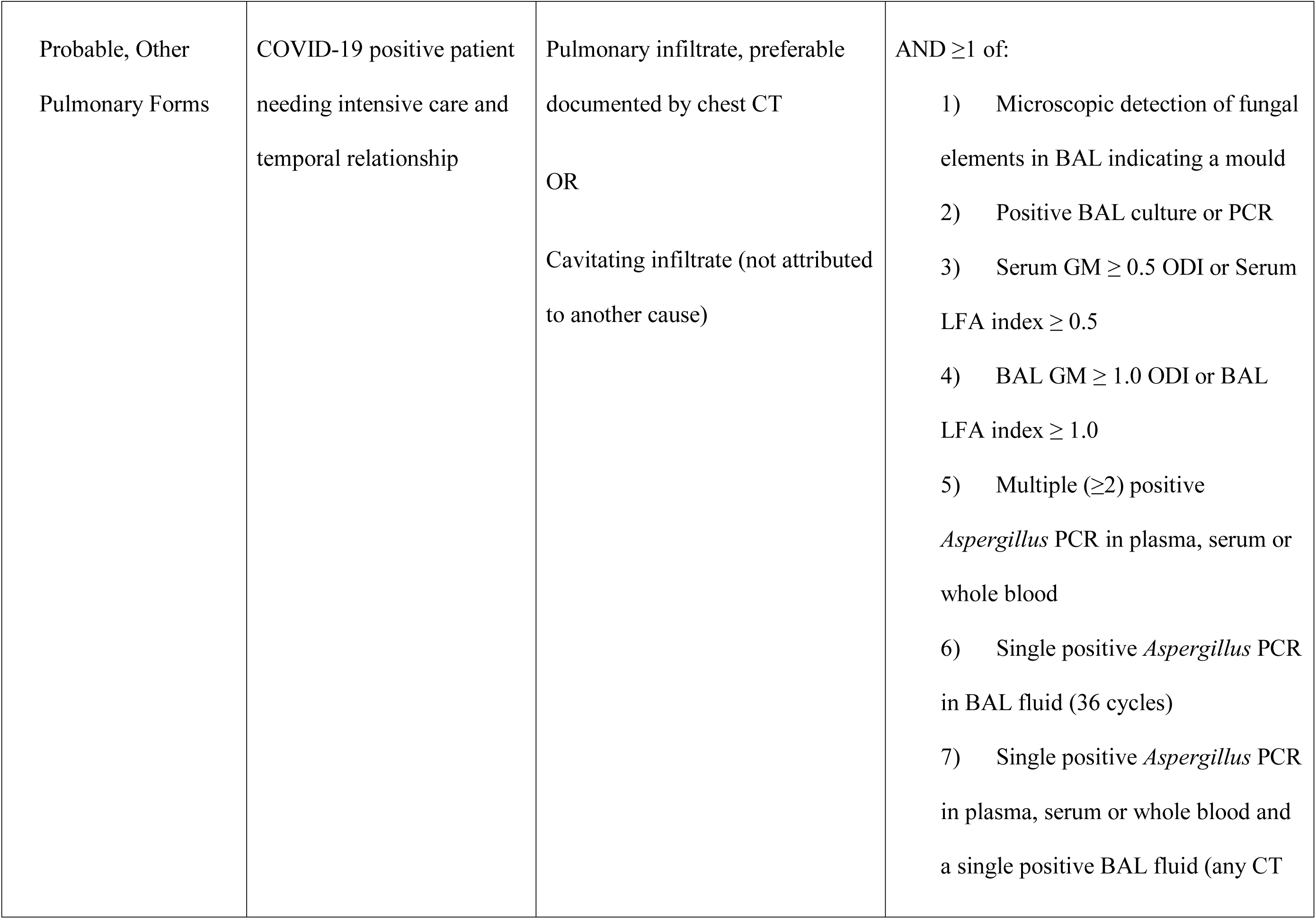
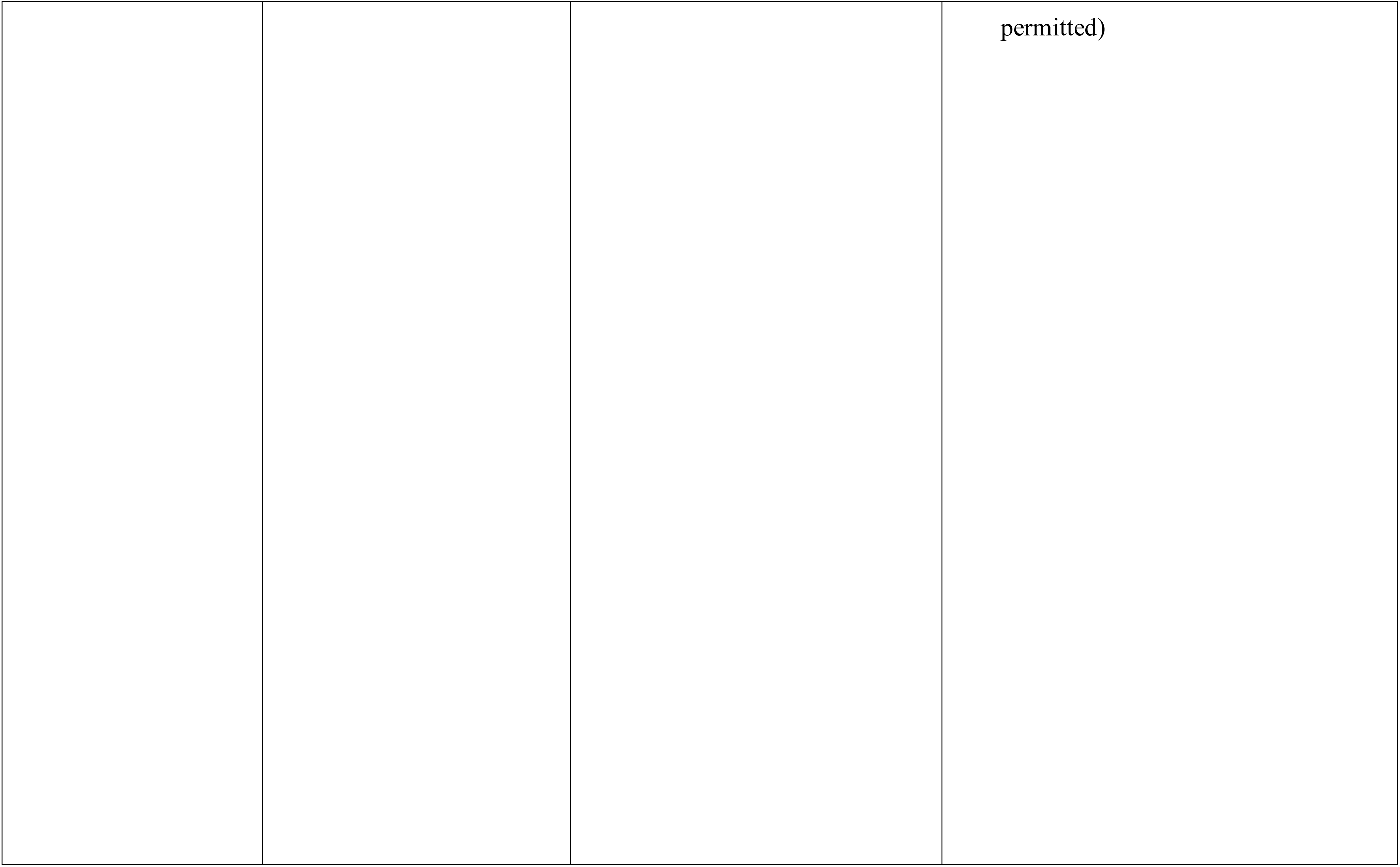
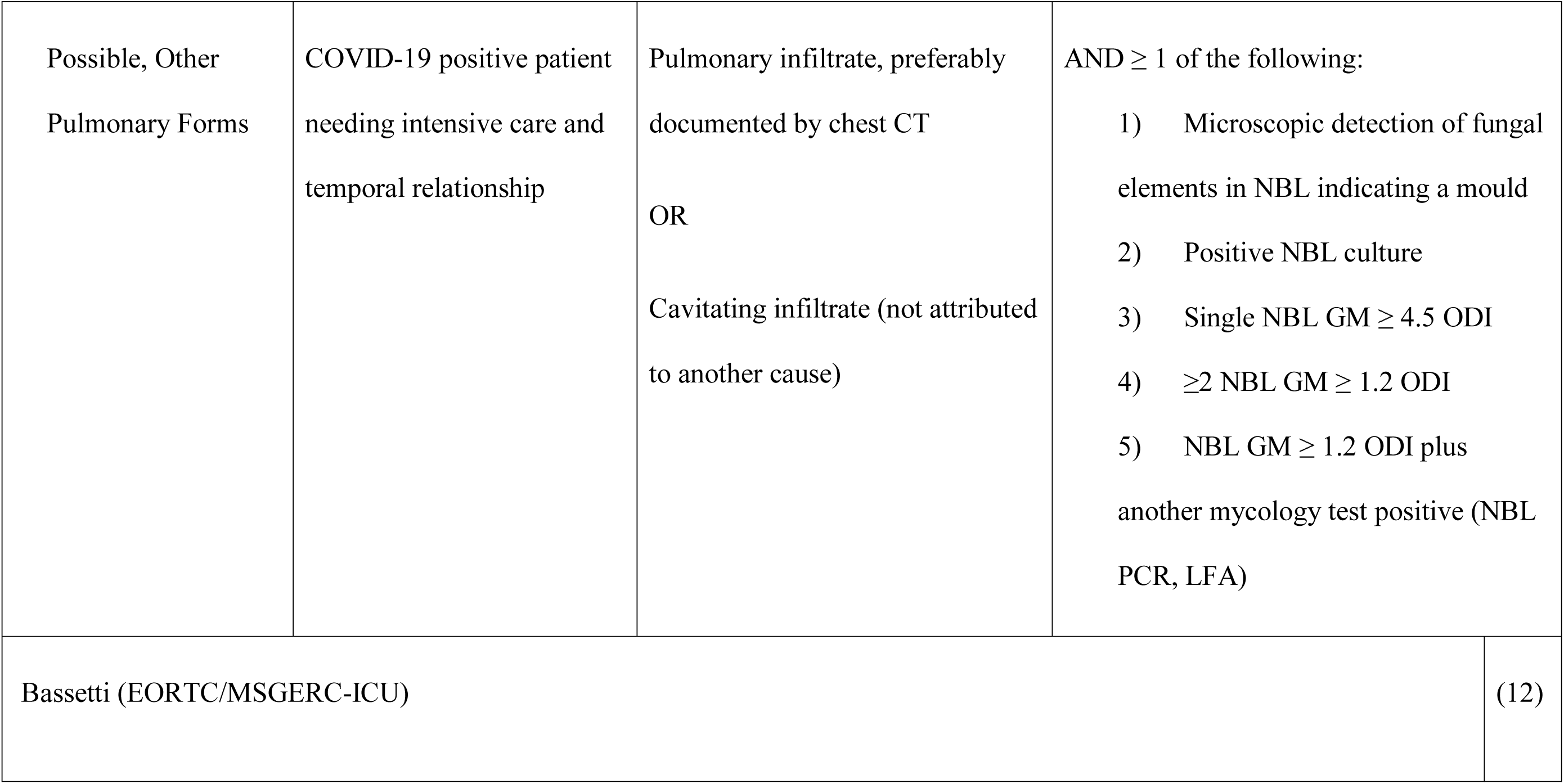
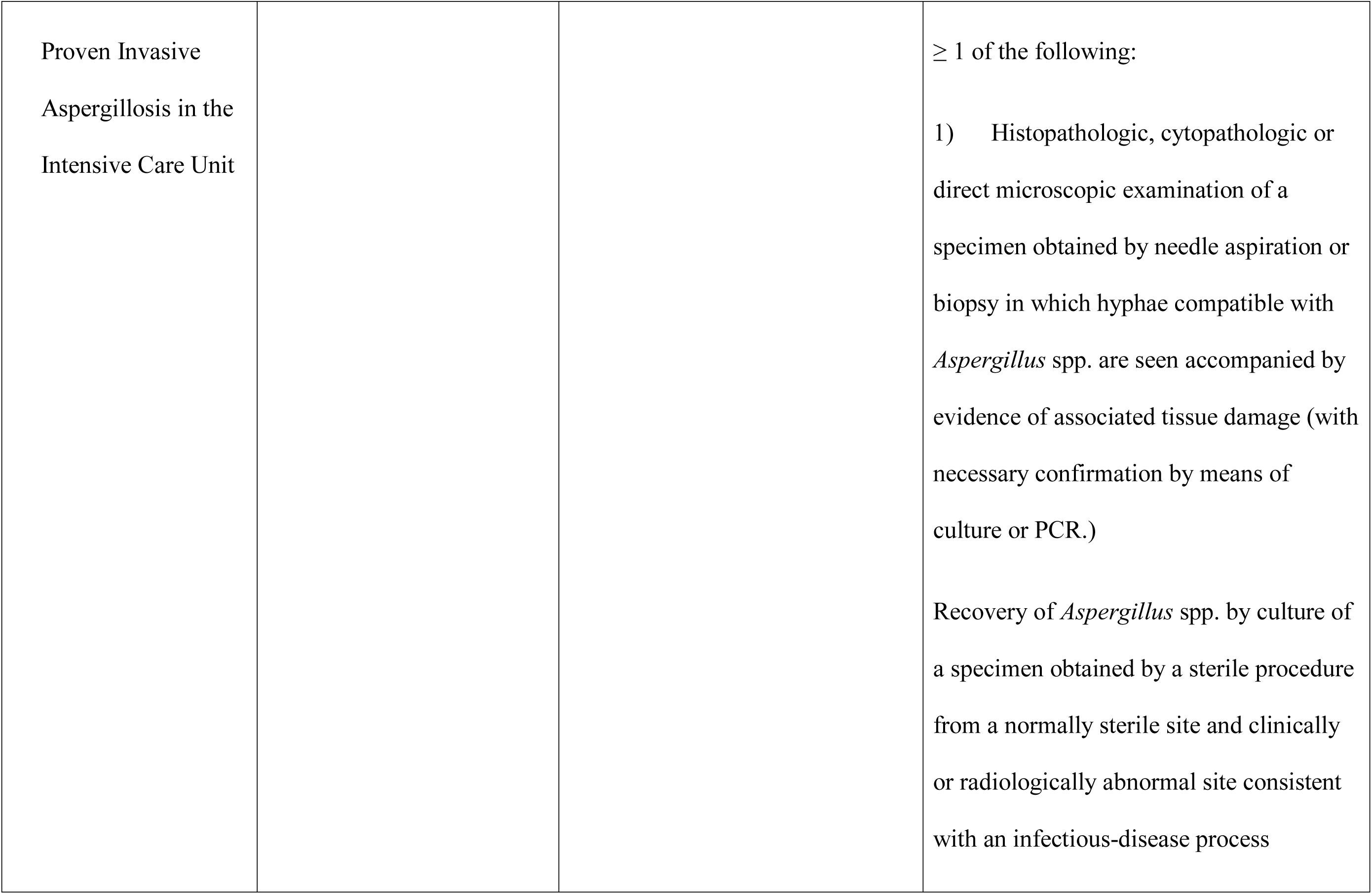
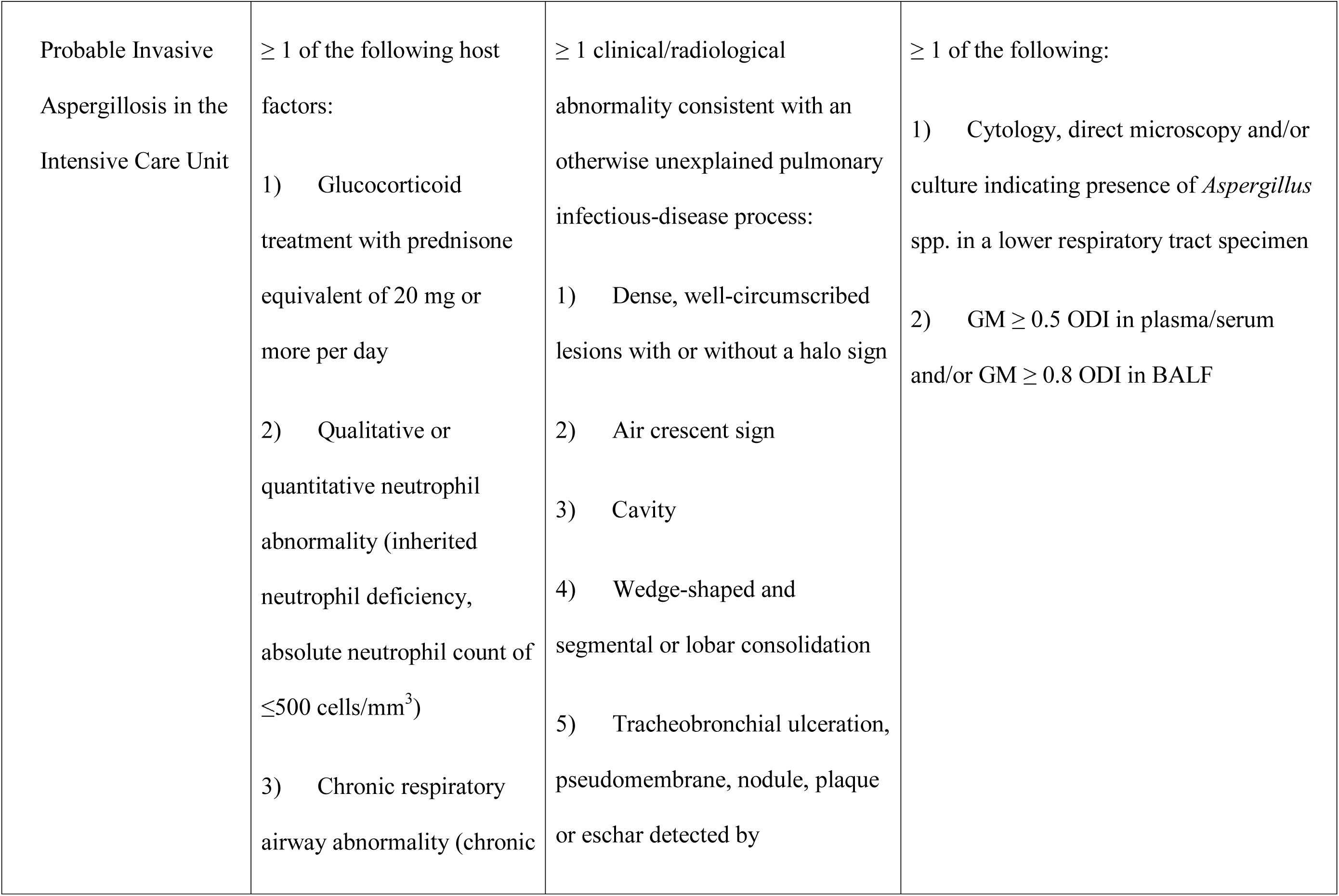
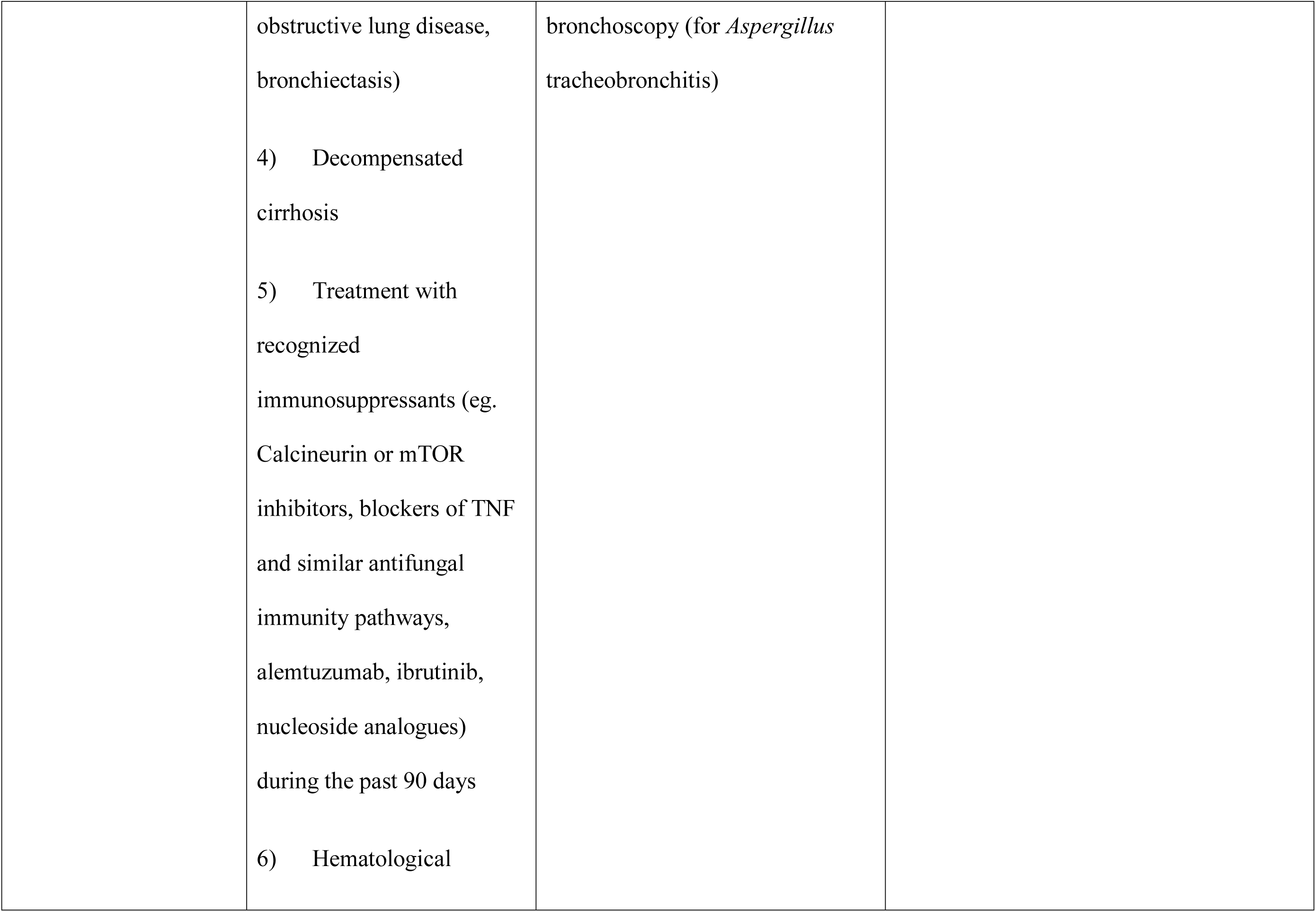
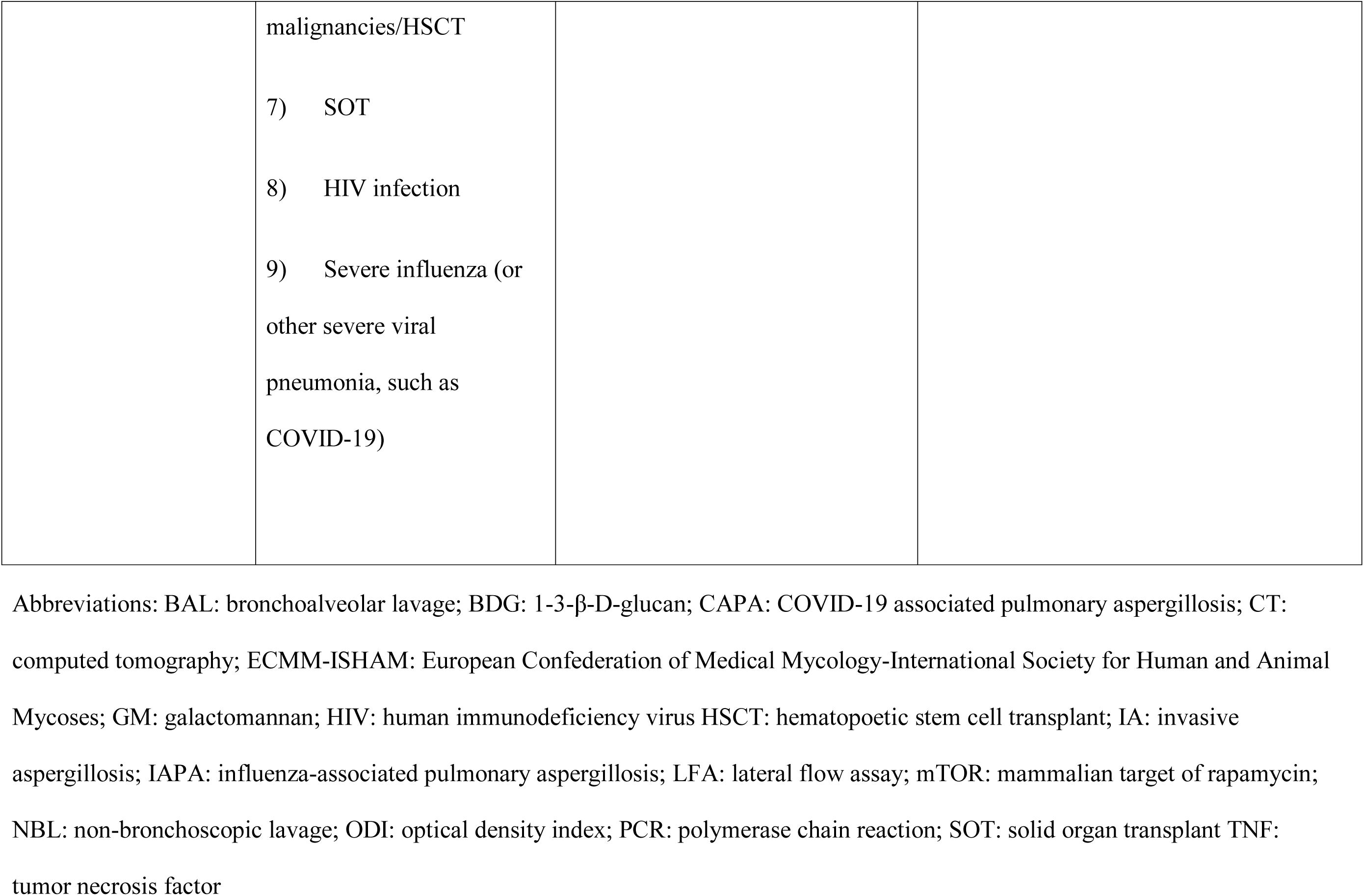
Research definitions of COVID-19 associated pulmonary aspergillosis

Given that some data was not always explicitly stated, we made some assumptions. Firstly, all patients admitted to an ICU with COVID-19 were assumed to meet the clinical criteria for the definition proposed by White et al, i.e. had at least 1 of: refractory fever despite greater than 3 days antibiotics; recrudescent fever of ≥48 hours despite antibiotics; dyspnea; hemoptysis; pleural rub or chest pain; worsening respiratory function despite antibiotics and ventilatory support. Secondly, where radiological findings were not provided, abnormal imaging was assumed for critically ill patients with COVID-19. Thirdly, for the purpose of White et al classification, radiographic abnormalities reported simply as being typical for COVID-19 were assumed to not be specific for pulmonary aspergillosis. On the other hand, because of the specific imaging requirements needed to meet the Bassetti et al classification, if radiographic results were not provided, these were considered unclassifiable. Immunocompromise was defined as per EORTC-MSGERC host factors for aspergillosis (8).

### Risk of Bias Quality Assessment

A formal assessment for risk of bias was conducted using an adaptation of the Joanna Briggs Institute (JBI) cohort checklist tool for systematic reviews (13). These tools rated the quality of study by assessing for appropriate patient population, fungal diagnostics, definitions, outcomes and/or follow-up. Two reviewers assessed the quality of all included studies and discrepancies were arbitrated by a third reviewer.

### Statistical Analysis

Descriptive data were reported for continuous outcomes. Dichotomous data and outcomes were reported as frequencies and proportions. The associations between antifungal therapy and mortality for patients with CAPA - defined as reported by authors and according to the 4 research definitions - were tested using Fisher’s exact test. We excluded from the “as reported” analysis studies that used a non-standard definition for CAPA that specifically excluded those patients who improved without therapy. Diagnostic definitions were compared with Spearman’s rank correlation (ρ), with p=0.05. Analyses were done using SPSS v26.0 (Armonk, New York, USA).

## Results

### Literature search

The search for publications identified 398 articles from PubMed (n=171), Embase (n=138), Web of Science (n=84), and grey literature and citations from general and systematic review papers (n=5). After removal of duplicates, 204 relevant studies were identified for title and abstract review. Eighty-three studies were eligible for full-text review, of which 38 met inclusion criteria, including 22 with patient-level data (Figure 1, Table 2). There were 35 cohort studies and 3 case series that were included. Individual patient details from studies meeting inclusion criteria are presented in Supplementary Table 1.

**Figure 1.**
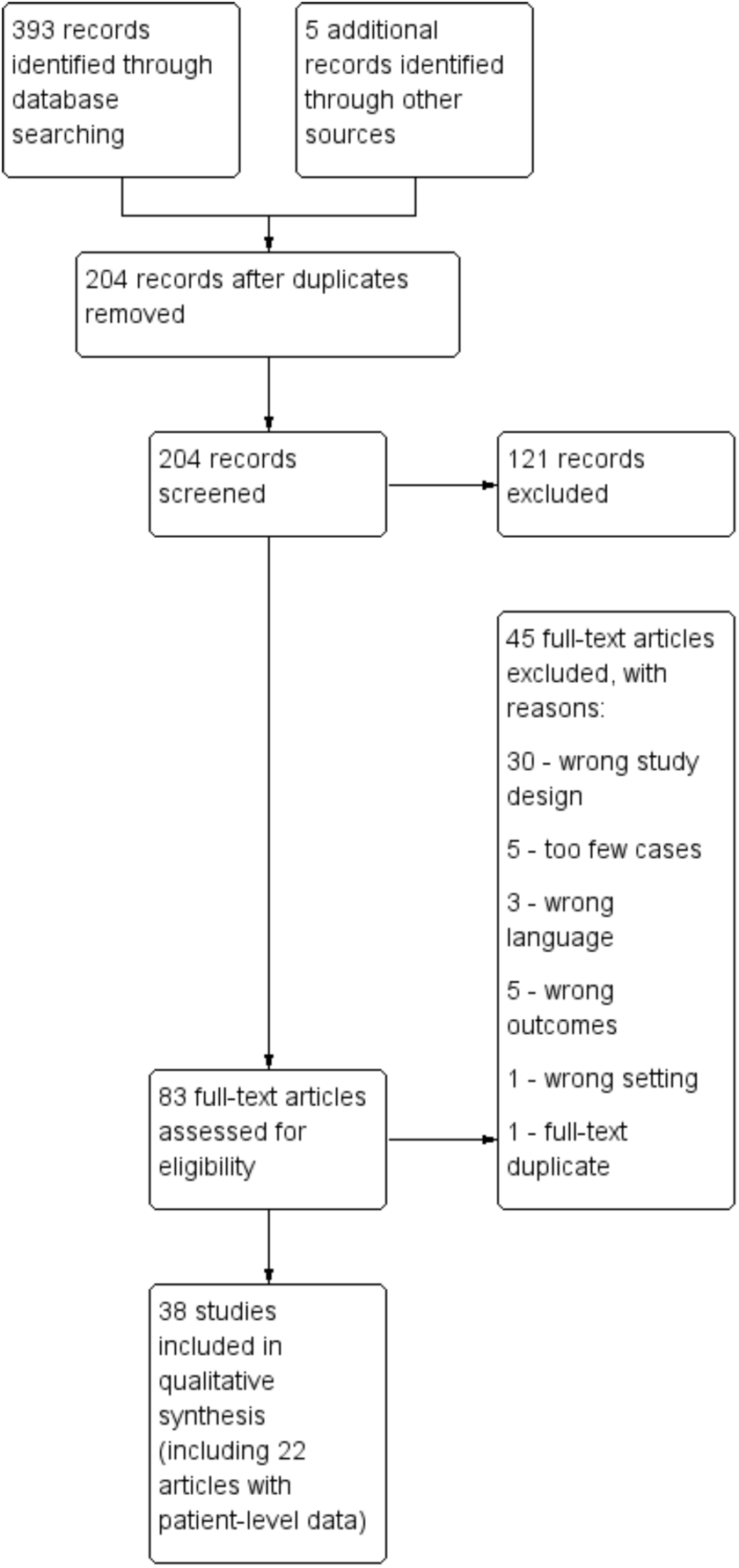
PRISMA flowchart.

**Table 2.**
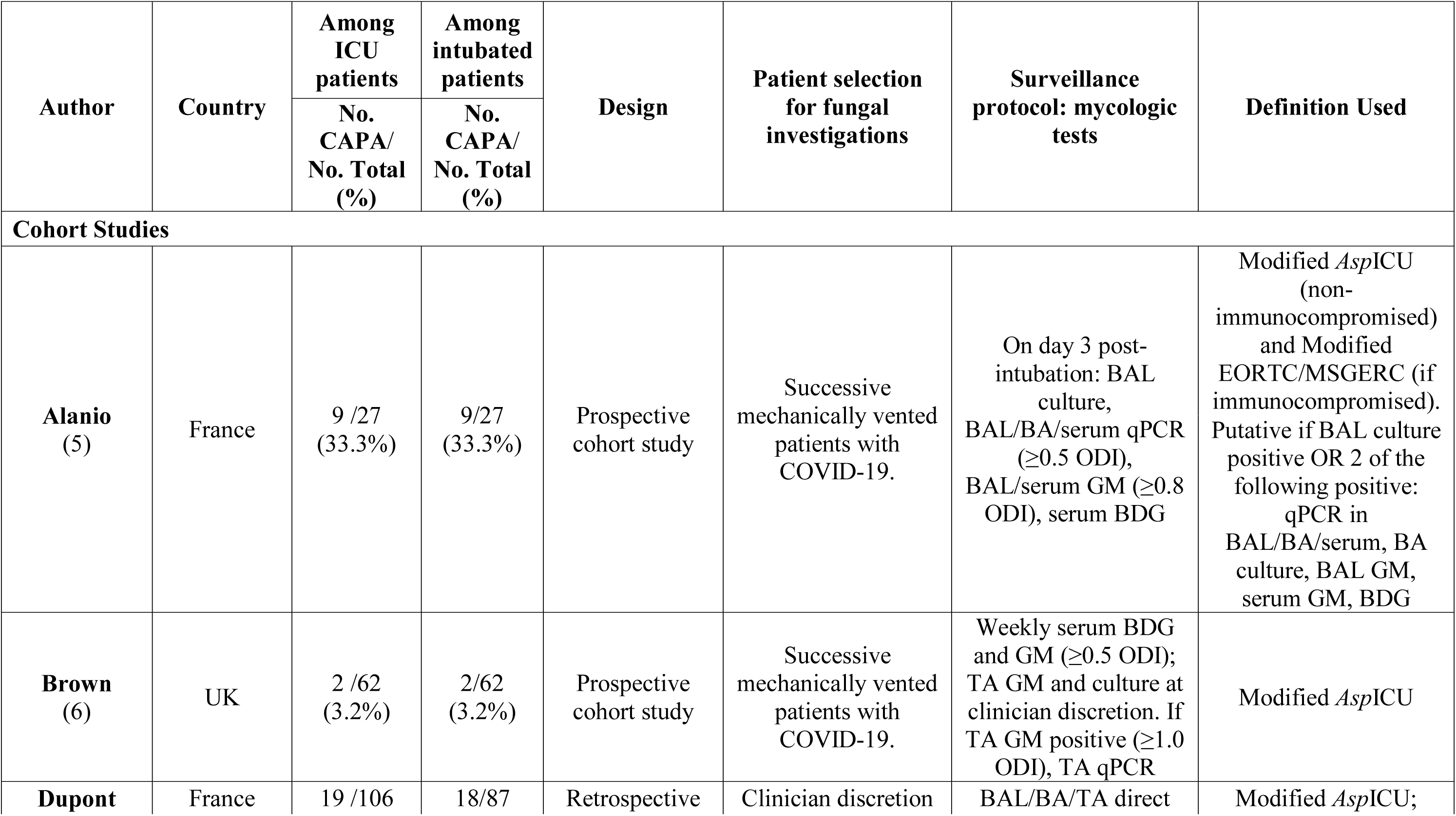
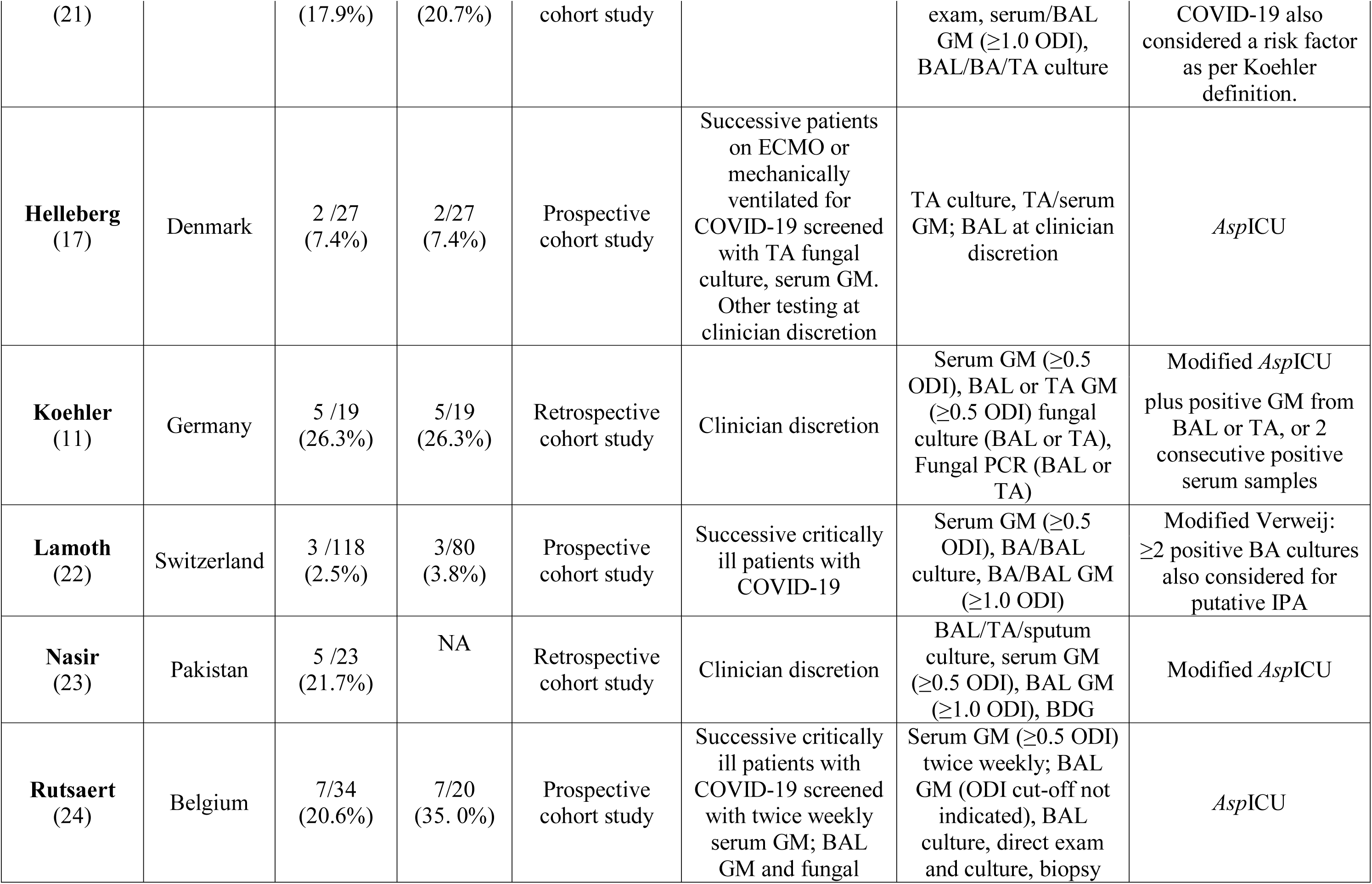
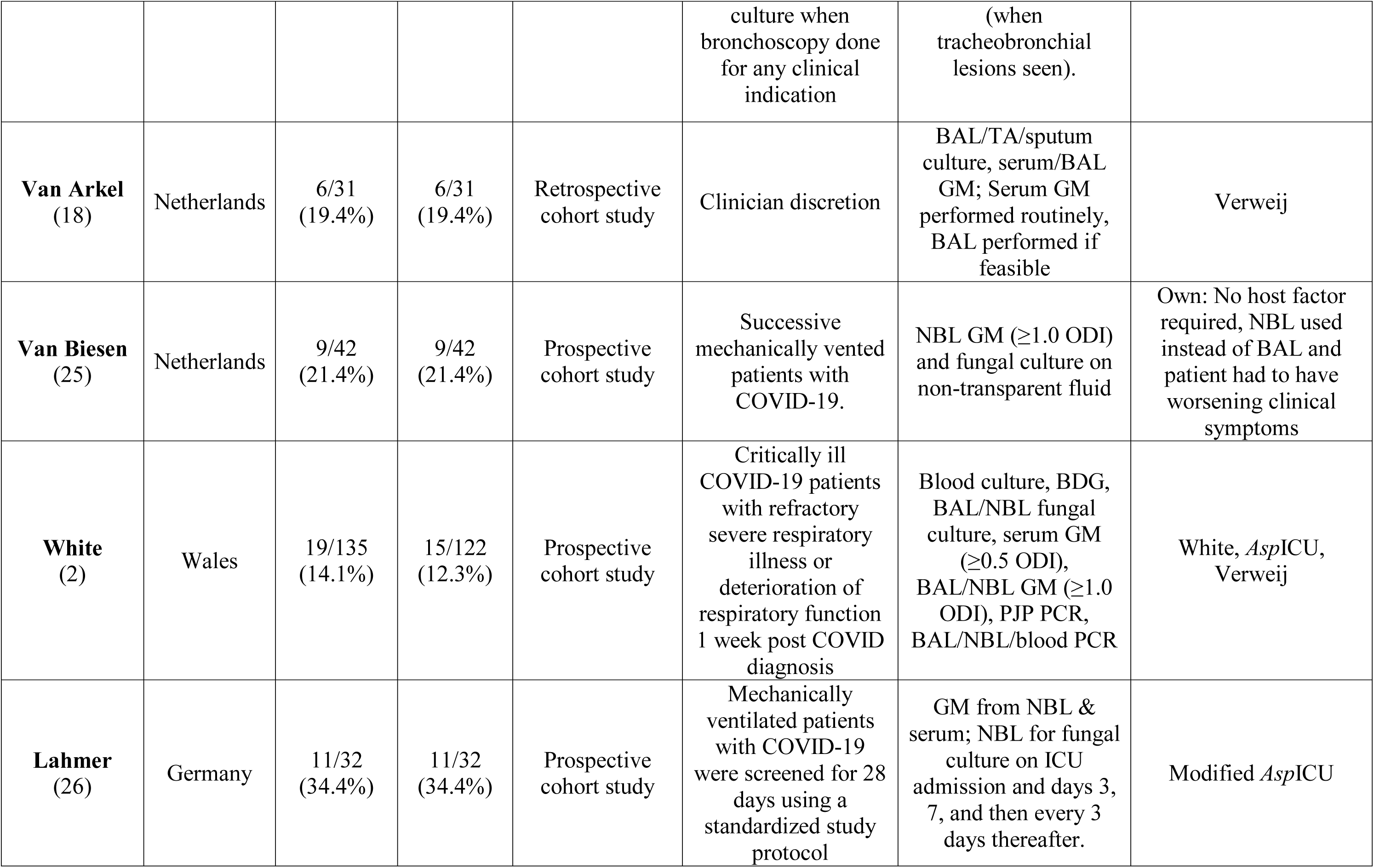
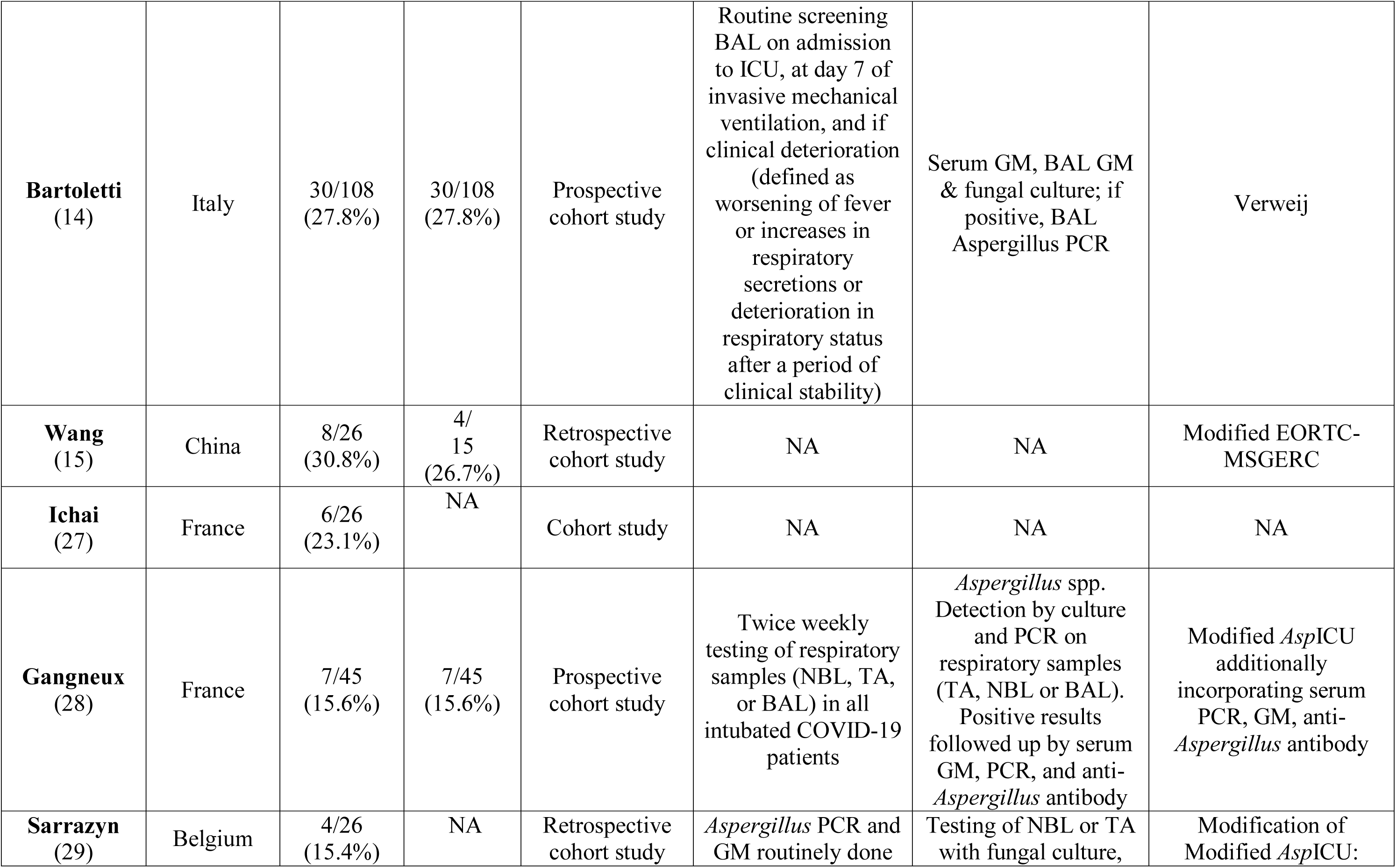
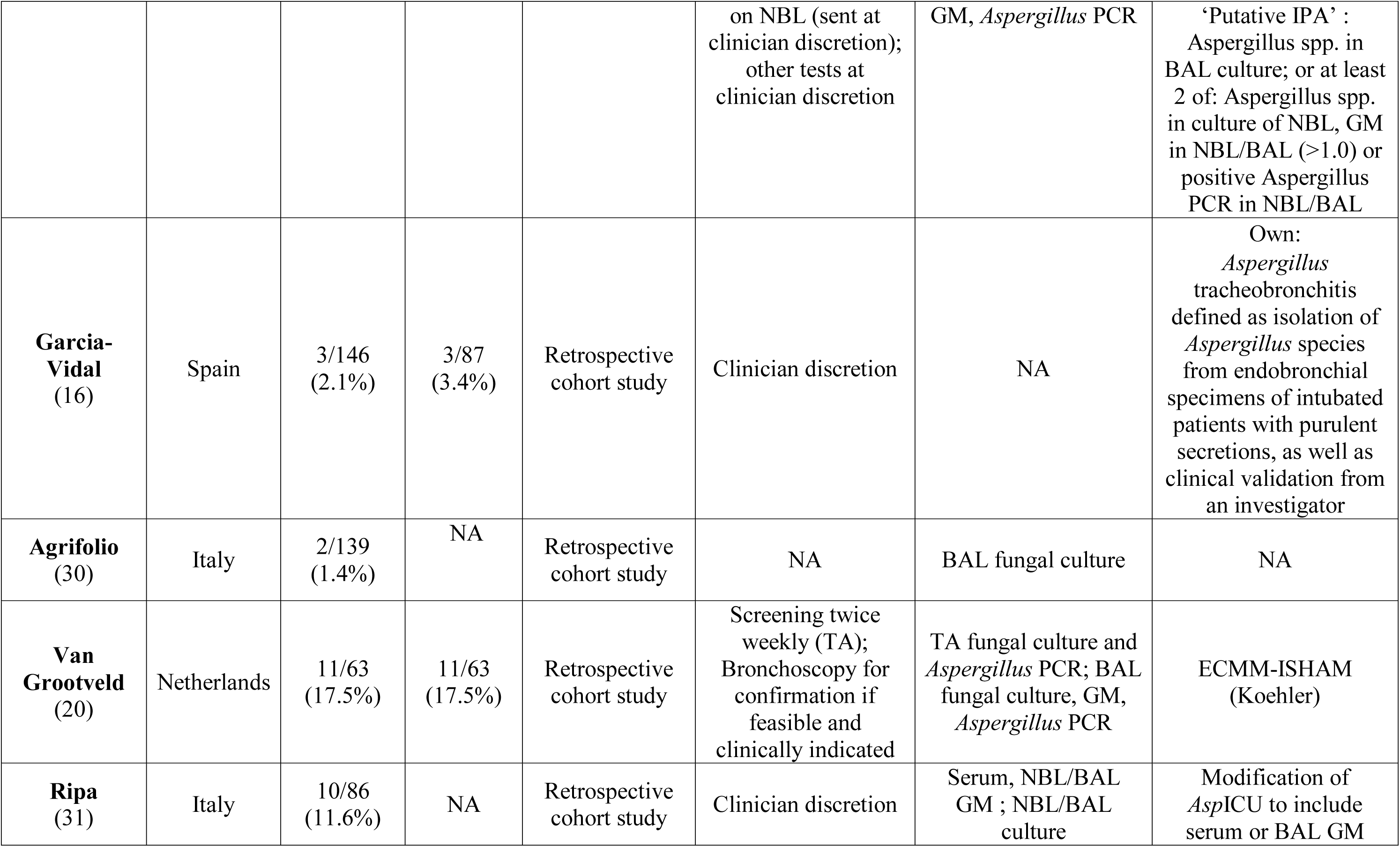
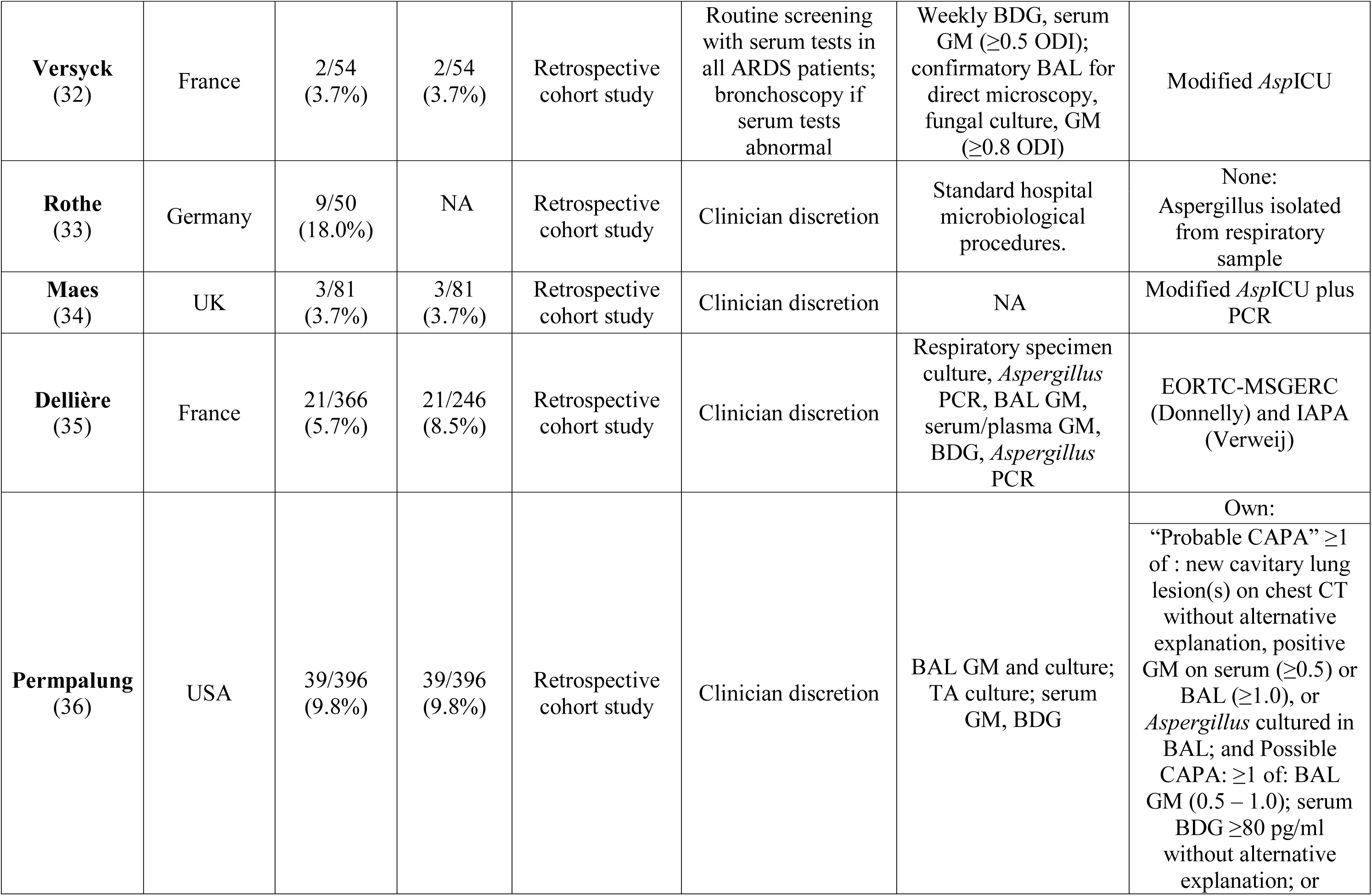
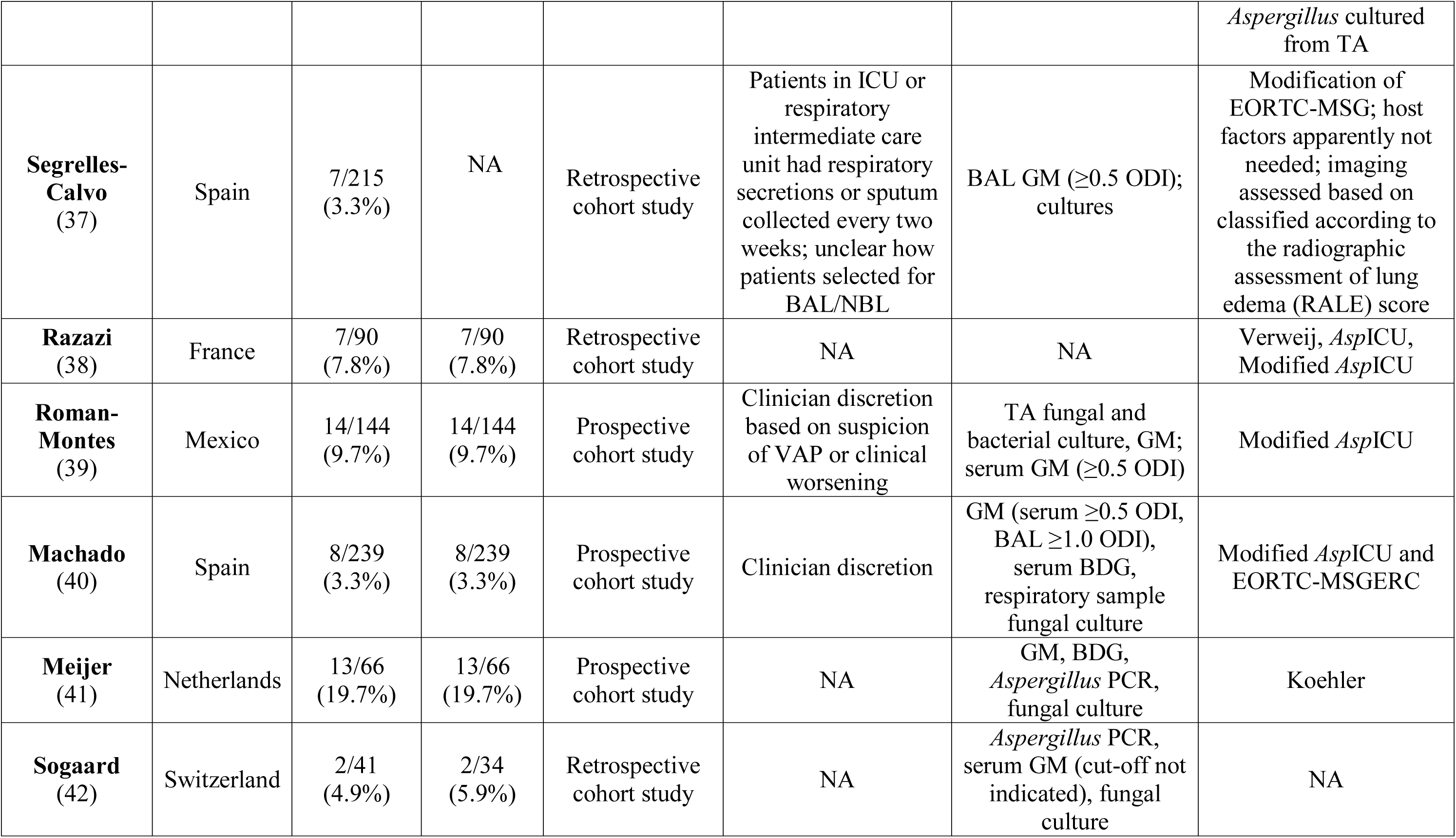
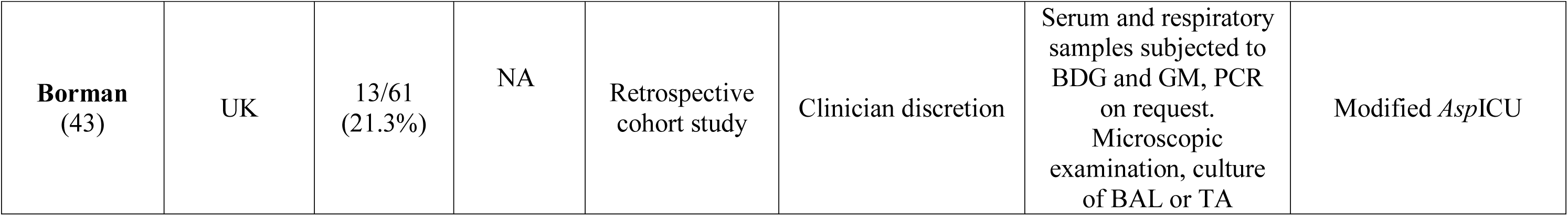
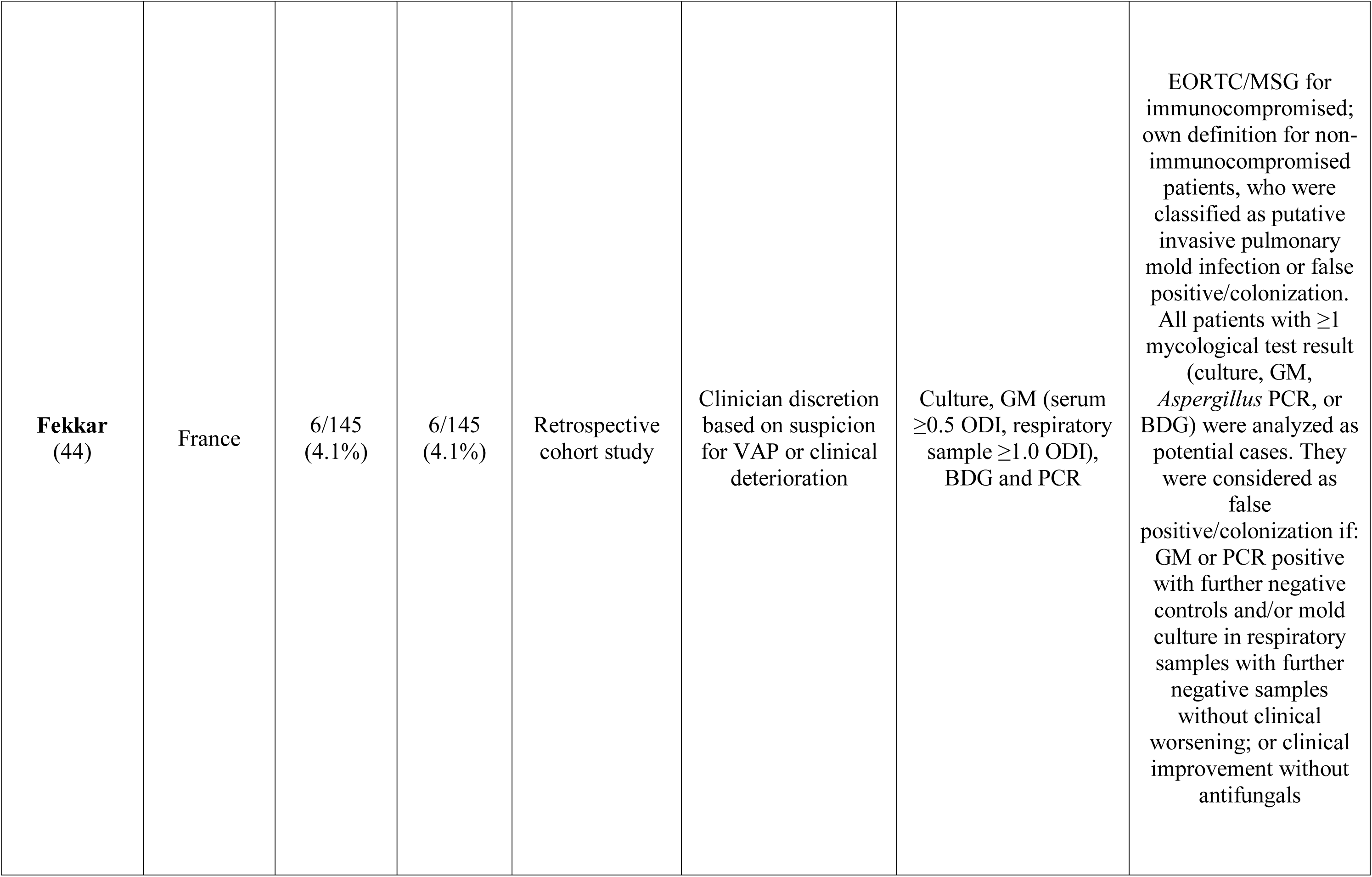
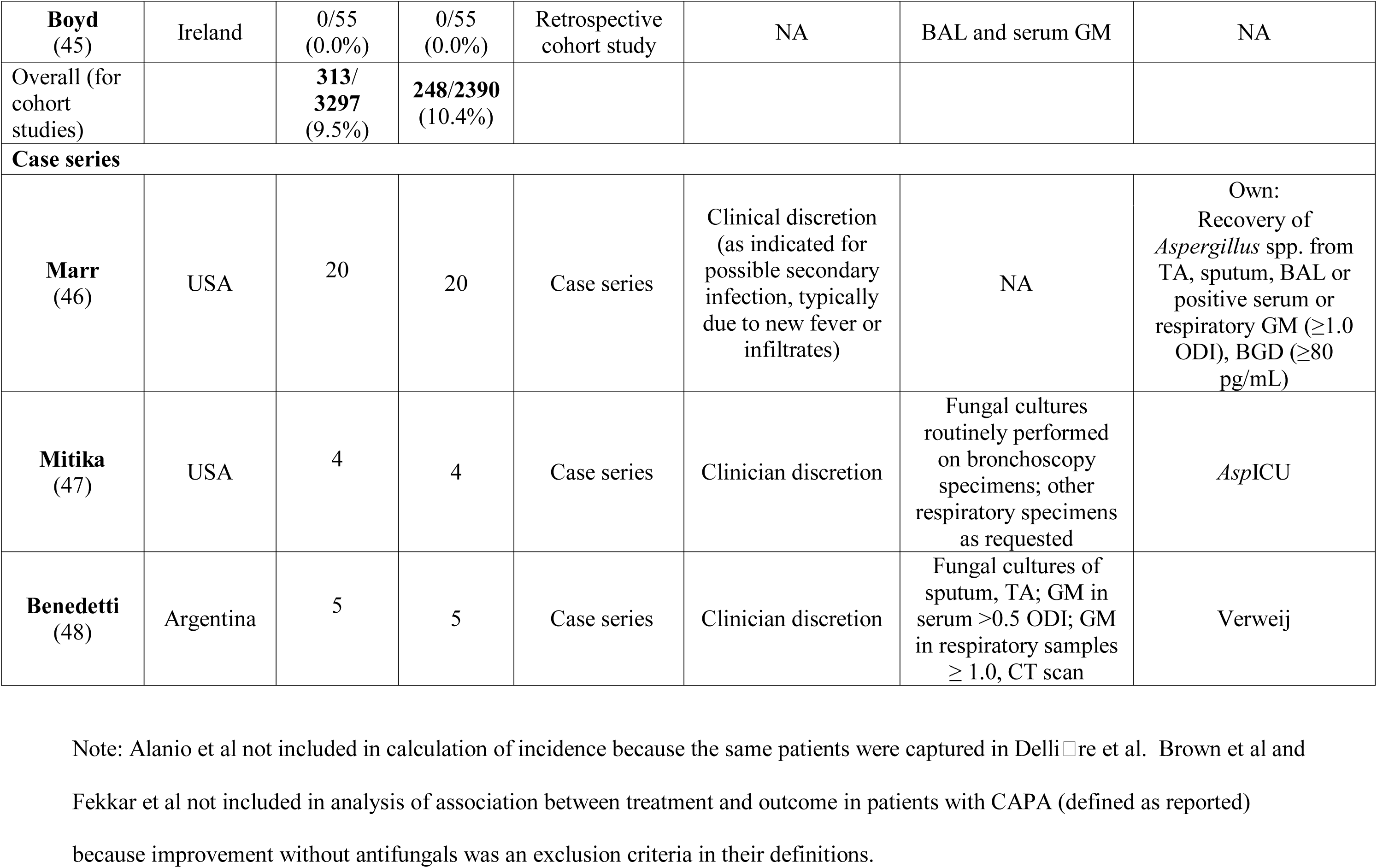
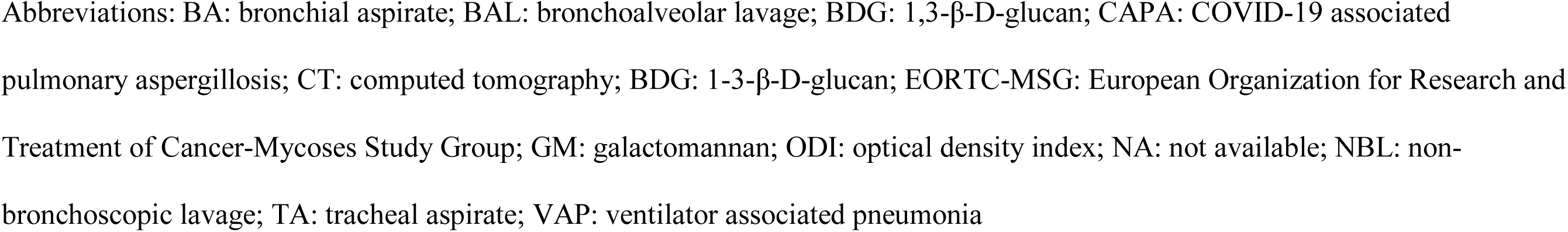
Characteristics and findings of included studies.

### Risk of Bias Assessment Quality Assessment

All studies met overall inclusion criteria with differences noted in Table 2. Given the low quality of evidence (non-RCT studies), a GRADE analysis could not be performed.

### Correlations between definitions

Four proposed definitions for CAPA were selected for analysis: (Verweij) (10), White (2), Koehler (11), Bassetti (12)), summarized in Table 1. From 22 studies, we identified 197 patients reported as having CAPA for whom patient level data was reported (Supplemental Table 1). Among these, 71 (36.0%) met criteria of the Verweij et al definitions (4 proven tracheobronchitis, and 67 probable, without documented tracheobronchitis), 79 (40.1%) met the White et al definitions (4 proven tracheobronchitis, 77 putative), 97 (49.2%) met the Koehler et al definitions (4 proven tracheobronchitis, 63 probable, other pulmonary forms, and 30 possible, other pulmonary forms). One-hundred-and-forty-five patients could be classified according to the Bassetti et al definitions, and 52 were unclassifiable due to lack of radiographic details. Among the 145 classifiable patients, 36 (24.8%) met the Bassetti et al criteria for CAPA (4 proven, 32 probable invasive aspergillosis). Seventy-six patients (38.6%) did not meet criteria for CAPA according to any definitions: 41 (20.8%) were classified as not meeting criteria for CAPA for all 4 definitions, and another 35 (17.8%) did not meet CAPA criteria for any of the Verweij, White, and Koehler definitions and were unclassifiable by Bassetti criteria. There was only modest agreement between any 2 definitions: the correlation coefficient, ρ, ranged between 0.330 and 0.621 (Table 2 and Figure 2).

**Figure 2.**
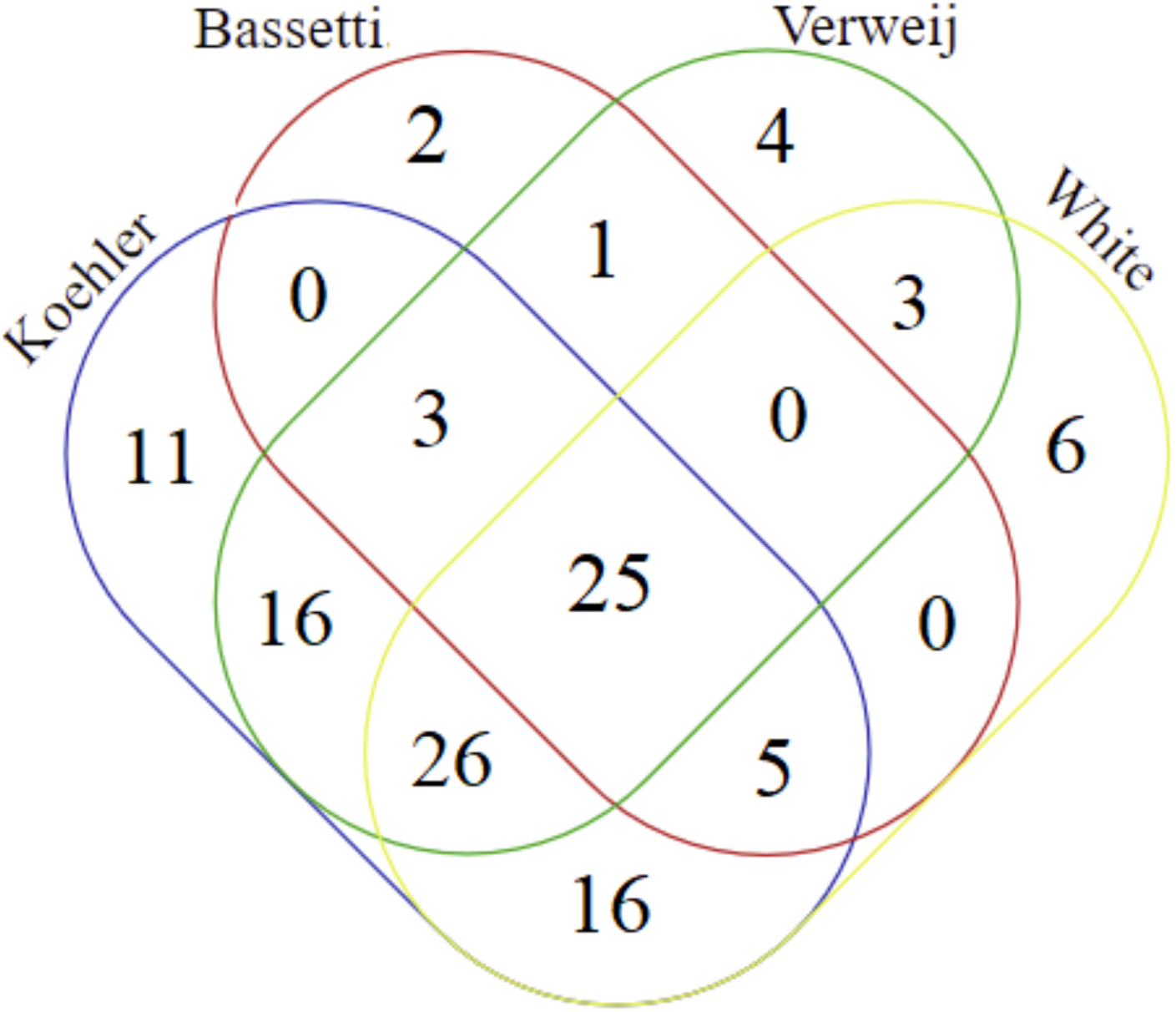
Venn diagram illustrating number of patients meeting each CAPA classification. Ninety-three (47%) additional patients reported in the literature as having CAPA did not meet *any* of these definitions. Fifty-two patients were unclassifiable per Bassetti criteria due to lack of radiographic details.

### Incidence of CAPA in the ICU

The incidence of CAPA in the ICU was calculated based on 3,297 patients across 34 cohort studies from 17 countries (Table 2). The incidence of CAPA in ICU patients ranged from 0-34.3% (Figure 3). Together, these studies reported 313 cases of CAPA, for a pooled incidence of 9.5% in ICU patients. Among 2,390 patients who were recorded as receiving invasive mechanical ventilation, there were 248 cases of CAPA reported, for a pooled incidence of 10.4%.

**Figure 3.**
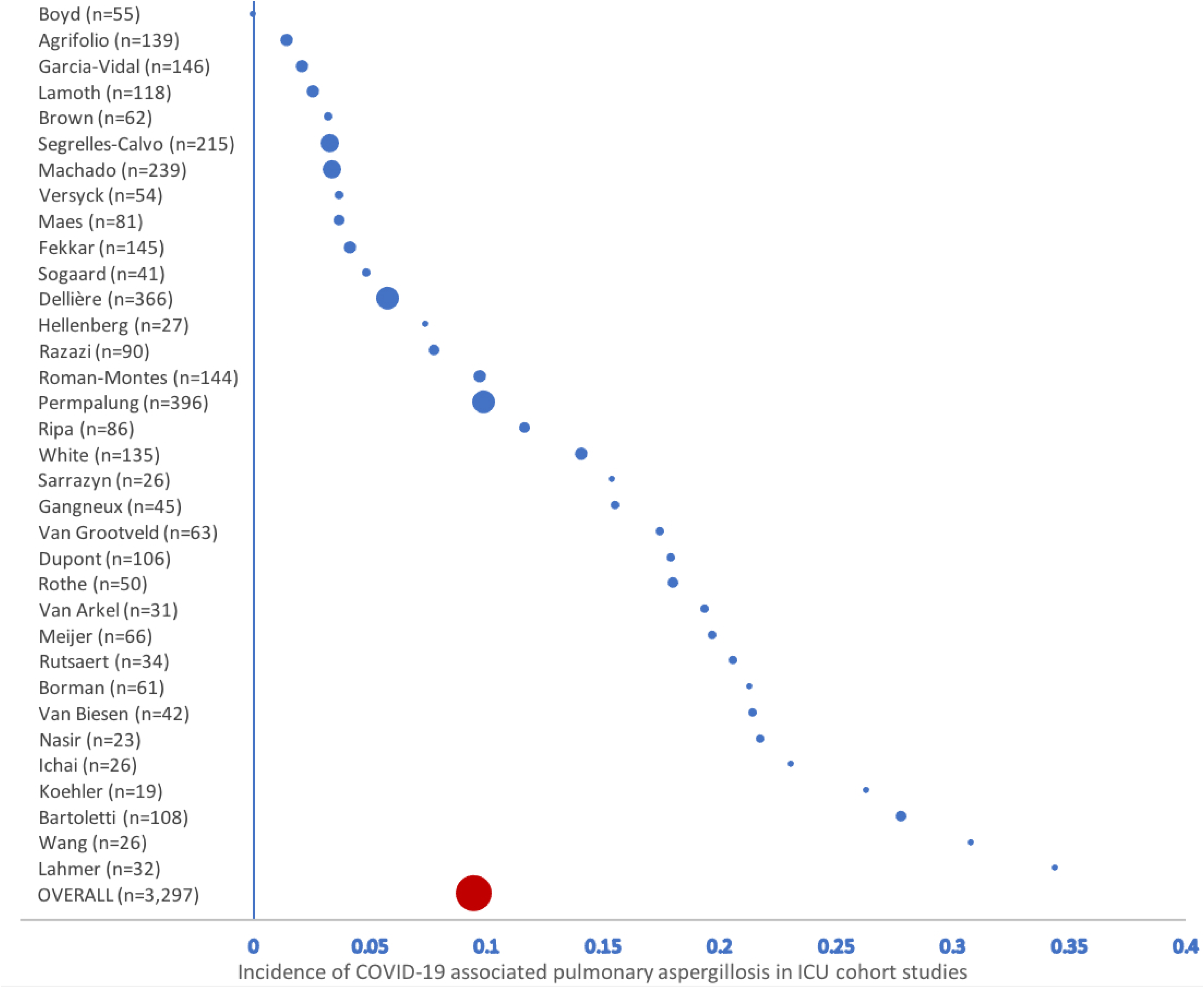
Reported incidence of COVID-19-associated pulmonary aspergillosis in ICU cohort studies.

Among only cohort studies with patient-level details (18 studies comprising 1,665 ICU patients), the pooled incidence of CAPA – as reported – was 9.2%; upon reclassification to Verweij, White, Koehler, and Bassetti definitions, the pooled incidence was 3.6%, 4.1%, 5.2% and 1.6%%, respectively (Table 4).

**Table 3.**
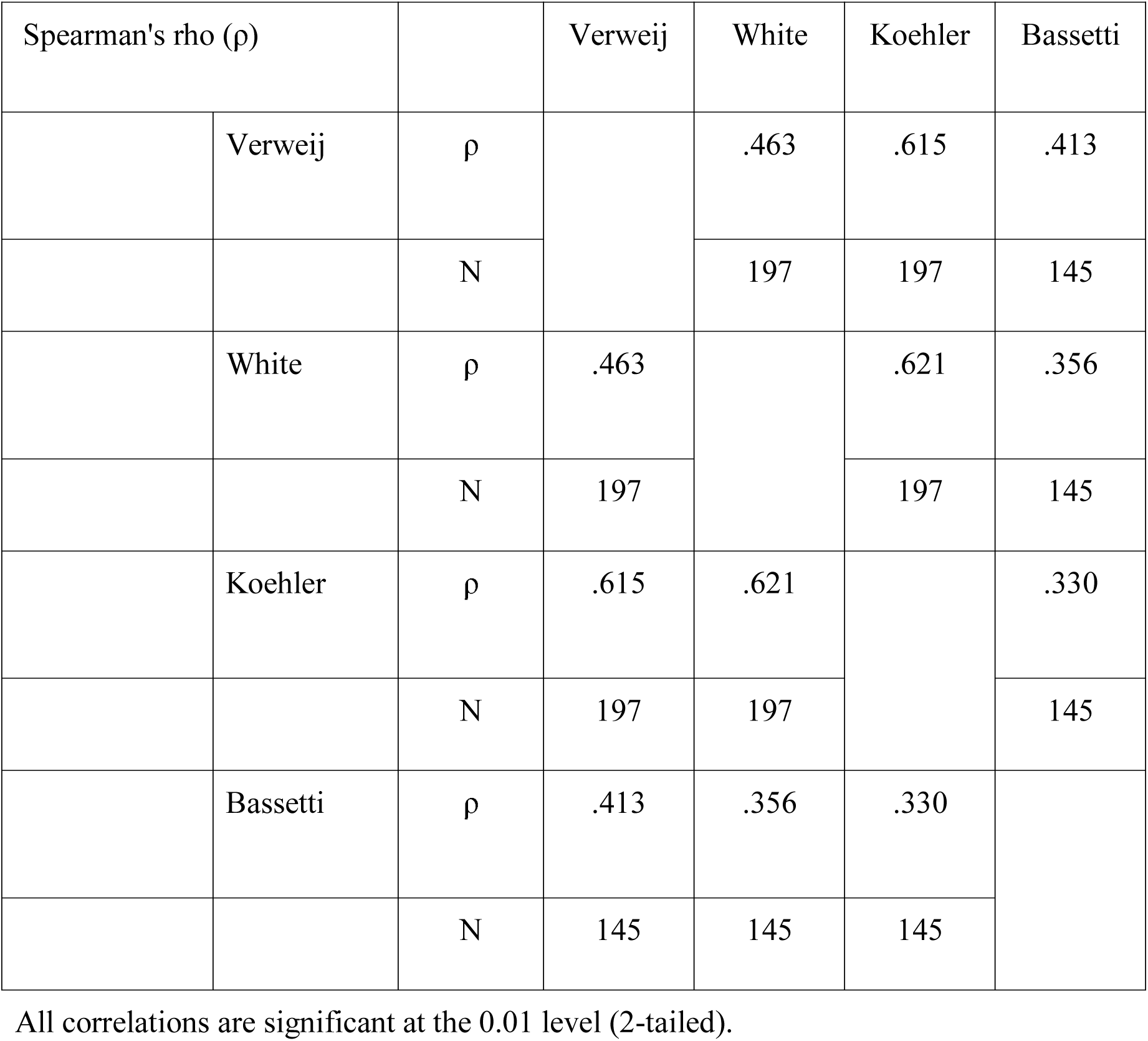
Correlations between 4 different definitions of COVID-19 associated pulmonary aspergillosis

**Table 4:** Incidence of CAPA in ICU cohort studies as reported and after reclassification according to 4 research definitions

| Study | Number ICU patients | Number of ICU patients with CAPA |  |  |  |  |
| --- | --- | --- | --- | --- | --- | --- |
|  |  | as reported | per Verweij | per White | per Koehler | per Bassetti |
| Alanio | 27 | 9 (33.3%) | 7 (25.9%) | 6 (22.2%) | 8 (29.6%) | 1 (11.1%) |
| Brown | 62 | 2 (3.2%) | 0 (0%) | 0 (0%) | 0 (0%) | 1 (1.6%)<sup>a</sup> |
| Dupont | 106 | 19 (17.9%) | 11 (10.4%) | 2 (1.9%) | 17 (16.0%) | 4 (3.8%) |
| Helleberg | 27 | 2 (7.4%) | 1 (3.7%) | 1 (3.7%) | 1 (3.7%) | 0 (0%) |
| Koehler | 19 | 5 (26.3%) | 4 (21.1%) | 4 (21.1%) | 4 (21.1%) | 5 (26.3%) |
| Lamoth | 118 | 3 (2.5%) | 1 (0.8%) | 3 (2.5%) | 3 (2.5%) | 0 (0%) |
| Nasir | 23 | 5 (21.7%) | 0 (0%) | 0 (0%) | 0 (0%) | 0 (0%) |
| Rutsaert | 34 | 7 (20.6%) | 6 (17.6%) | 5 (14.7%) | 6 (17.6%) | 4 (11.8%)<sup>b</sup> |
| Van Arkel | 31 | 6 (19.4%) | 3 (9.7%) | 1 (3.2%) | 1 (3.2%) | 0 (0%)<sup>c</sup> |
| Van Biesen | 42 | 9 (21.4%) | 0 (0%) | 7 (16.7%) | 7 (16.7%) | 0 (0%) |
| White | 135 | 25 (18.5%) | 3 (2.2%) | 19 (14.1%) | 15 (11.1%) | 5 (3.7%) |
| Van<br>Grootveld | 63 | 11 (17.5%) | 10 (15.9%) | 8 (12.7%) | 9 (14.3%) | 1 (1.6%) |
| Versyck | 54 | 2 (3.7%) | 2 (3.7%) | 2 (3.7%) | 2 (3.7%) | 2 (3.7%) |
| Segrelles-Calvo | 215 | 7 (3.3%) | 0 (0%) | 0 (0%) | 0 (0%) | 0 (0%) |
| Roman-Montes | 144 | 14 (9.7%) | 6 (4.2%) | 5 (83.3%) | 6 (4.2%) | 0 (0%) |
| Machado | 239 | 8 (3.3%) | 3 (1.3%) | 3 (1.3%) | 3 (1.3%) | 1 (0.4%)<sup>b</sup> |
| Fekkar | 260 | 6 (2.3%) | 0 (0%) | 0 (0%) | 0 (0%) | 0 (0%)<sup>c</sup> |
| Meijer | 66 | 13 (19.7%) | 3 (4.5%) | 3 (4.5%) | 4 (6.1%) | 2 (3.0%) |
| Pooled<br>Total | 1,665 | 153 (9.2%) | 60 (3.6%) | 69 (4.1%) | 86 (5.2%) | 26 (1.6%) |

Note: Only ICU cohort studies that included patient-level details allowing CAPA reclassification are included. Abbreviations: CAPA – COVID-19-associated pulmonary aspergillosis; ICU – intensive care unit. Footnotes: <sup>a</sup>1 patient was unclassifiable; <sup>b</sup>3 patients were unclassifiable; <sup>c</sup>6 unclassifiable;

### Patient Characteristics and Diagnosis

Twenty-two articles reporting 197 individuals with CAPA were included in the patient-level analysis (Supplementary Table 1). Demographic and clinical features of these cases are summarized in Table 5. The median age was 64 years (IQR 55 – 73) and 72.7% were male. Immunocompromise status was present in 9 patients (4.6%). Eighty-one patients received immunomodulatory therapy: 67 patients (33.5%) received corticosteroids (in addition to 4 patients already on steroids at admission), and 29 (14.7%) received tocilizumab (15 patients received both). Additionally, patients were treated with the following therapies, alone or in combination: azithromycin (n=19), chloroquine or hydroxychloroquine (n=43), lopinavir/ritonavir or darunavir/cobicistat (n=24), ribavirin (n=1), interferon-β1b (n=5), and remdesivir (n=5).

**Table 5.** Demographic and clinical features reported for 197 patients reported to have CAPA for whom patient-level details are available

| Characteristic |  |
| --- | --- |
| Age, Median years (IQR) | 64 (55 – 73)<sup>a</sup> |
| Sex, Males (%) | 125/172 (72.7%) |
| Immune compromised* | 9 (5%) |
| Systemic steroids | 66 (33.5%)** |
| Tocilizumab | 28 (14.2%) |
| Azithromycin | 19 (9.6%) |
| Invasive mechanical ventilation | 149 (84.2%) |
| ECMO | 7 (3.6%) |
| Radiography |  |
| Nodules | 25*** |
| Cavitations | 17 |
| Bronchoscopy | 76/155 (38.6%) |
| Tracheobronchial abnormalities**** | 4/76 (5.3%) |
| Mould-active antifungals | 141/182 (77.5%) |
| Deaths | 91/182 (50.0%) |
| Timing of CAPA diagnosis, median days from ICU admission (IQR)<sup>b</sup> | 9 (5 - 14)<sup>b</sup> |
| Timing of CAPA diagnosis, median days of mechanical ventilation before diagnosis (IQR) | 6 (3-10)<sup>c</sup> |

Data missing for: <sup>a</sup>25 patients, <sup>b</sup>124 patients, <sup>c</sup>108 patients
Notes: \* Per European Organization for Research and Treatment of Cancer – Mycoses Study Group Education and Research Consortium consensus definitions for host factors; \*\*plus 4 already on steroids; \*\*\*1 additional patient had nodules attributed to known pulmonary metastases.
Abbreviations: CAPA: COVID-19 associated pulmonary aspergillosis; ICU: intensive care unit; IQR: interquartile range; ECMO: extra-corporeal membrane oxygenation

One-hundred-and-forty-nine patients (84.2%) received invasive mechanical ventilation, and 7 (3.6%) received extracorporeal membrane oxygenation (ECMO). The duration of invasive mechanical ventilation at CAPA diagnosis, known for 41 patients (27.5%), was a median of 6 days (IQR 3-10); duration of ICU admission at time of CAPA diagnosis, known for 73 patients (37.1%), was a median of 9 days (IQR 5-14).

Radiographic findings were described for 142 patients (72.1%). These were reported as showing non-specific abnormalities (n=13), infiltrates (n=24), consolidation (n=36), nodules (n=25), cavitations (n=17), and ground-glass opacities (n=28), or they were simply reported as being typical for COVID-19 (n=42).

Among patients reported to have CAPA, bronchoscopy was performed on 80 (40.6%), among whom just 4 (5.3%) – all from a single report – had tracheobronchial abnormalities, and in these patients tracheobronchial biopsies had histopathological evidence of invasive aspergillosis. At least 8 additional patients from cohort studies that lacked patient-level details had lesions on bronchoscopy (14, 15). Bartoletti et al reported routine bronchoscopies on ICU admission, at 7 days, and with clinical deterioration (14). Thirty out of 108 patients were diagnosed with CAPA, including 6 (20%) with endobronchial lesions visualized on bronchoscopy.

Bronchial sampling was done on 123 CAPA patients, including bronchoscopic (n=80) and non-bronchoscopic lavage (NBL) (n=43). One-hundred sixteen patients had *Aspergillus* spp. cultured from a respiratory specimen. These included: *A. fumigatus* in 98 (84.5%), *A. flavus* in 4 (3.4%), *A niger* in 2 (1.7%), *A. citronotterreus, A. lentelus, A. versicolor, Aspergillus* section fumigati*, A. terreus* and *A. calidoustus* in 1 (0.8%) each and 1 unspeciated *Aspergillus* spp. Mixed infections were encountered in 5 (2.6%) additional patients: 3 with *A. fumigatus* and *A. flavus,* 1 with *A. fumigatus* with *A. versicolor* and 1 with *A, fumigatus, A. awamori* and *A. terreus.* These were cultured from BAL (n=45), bronchial aspirates (n=17), NBL (n=18), endotracheal aspirates (n=34), and sputum (n=2). PCR for *Aspergillus* species was done in 81 patients (41.1%) and was positive in 47 of these (58.0%). These included from BAL/BA in 9 patients, from NBL in 10 patients, from NBL and plasma in 2 patients, from NBL and serum in 2 patients, from bronchial aspirate in patients; from tracheal aspirate (TA) in 5 patients, from TA and BAL/BA in 6 patients. Four had PCR detection of *Aspergillus* species in serum, 1 from plasma and 1 from sputum.

Galactomannan was measured on bronchial fluid for 91 (46.2%) patients; 66 (72.5%) of these were positive (optical density index [ODI] ≥1) (Supplementary Table 1). Serum galactomannan was conducted on 81 (41.1%) patients, with 31 (38.3%) being positive (ODI ≥ 0.5). Serum BDG was conducted on 71 (36.0%) patients, of whom 36 (50.7%) had levels ≥80 pg/mL. Direct observation of branching hyphae was only reported in 3 patients (1.5%).

### Treatment and Outcomes

Treatment and survival data was available for 182 patients. One-hundred-forty-one patients (77.5%) were treated with mould-active antifungals, including (alone or in combination) a mould-active azole (voriconazole, itraconazole, isavuconazole, or posaconazole; n=110), amphotericin B (n=34), an echinocandin (n=22; used with a mould-active azole or amphotericin B in 20 cases, and alone in 2 cases); in 18 patients, the mould-active antifungal used was unclear. Forty-one (22.5%) patients did not receive mould-active antifungals.

In total, 91 patients survived (50.0%) and 91 patients died (50.0%). Among patients treated with mould-active antifungals, the survival rate was 53.0% (71/134) vs 41.4% (17/41) for patients who were not treated with mould-active antifungals (p=0.28). The lack of association between antifungal treatment and outcome persisted when the analysis was repeated for just those meeting each of the 4 CAPA definitions (0.37 < p < 1.0) (Table 6).

**Table 6.** Comparison of antifungal treatment and survival across different COVID-19 associated pulmonary aspergillosis (CAPA) definitions

| **CAPA definition** | **Survived/Treated (%)** | **Survived/Not treated (%)** | ***P*-value** |
| --- | --- | --- | --- |
| **As reported (n=174)** | 71/134 (52.9%) | 17/40 (41.4%) | 0.28 |
| **Verweij (n=70)** | 25/54 (46.3%) | 9/16 (56.3%) | 0.57 |
| **White (n=77)** | 34/64 (53.1%) | 5/13 (38.5%) | 0.37 |
| **Koehler (n=96)** | 41/73 (56.2%) | 12/23 (52.2%) | 0.81 |
| **Koehler**<br>**(proven/probable only)**<br>**(n=66)** | 24/51 (47.1%) | 8/15 (53.3%) | 0.77 |
| **Bassetti (n=34)** | 17/30 (56.7%) | 2/4 (50.0%) | 1.0 |

Fisher's exact test with two-tailed *P* values.

## Discussion

In this systematic review, we reviewed the incidence, diagnosis, management, and outcomes of CAPA, and compared research definitions. Overall, we found that the pooled incidence of CAPA, as reported, was 9.5% and varied widely between studies (0.0% to 34.3%), which may relate in part to surveillance practices and definitions used in individual reports.

Most patients with CAPA received invasive mechanical ventilation, although this association may be due to easier access to the lower respiratory tree for sampling and diagnosis. Most studies did not report bronchoscopy rates across the ICU cohort, so it is difficult to know if CAPA rates reflected discrepant use of this procedure in different centres. A small proportion of patients (5.6%) who had bronchoscopies were reported to have tracheobronchial abnormalities visualized. This is in contrast to IAPA, which has been observed to involve the tracheobronchial tree in up to half of all cases (10). While the presence of tracheobronchial disease should increase suspicion for CAPA, radiographic findings are insufficient to distinguish patients with CAPA from those with COVID-19 alone. In fact, a small minority of CAPA patients were reported to have pulmonary nodules or cavitations, findings classically associated with invasive aspergillosis in immunocompromised hosts.

*Aspergillus* spp. were most commonly detected through bronchial fluid fungal culture and, to a lesser extent, galactomannan or PCR. A significant minority of patients had bronchial fluid sampling by NBL. The precise role for NBL in diagnosing CAPA is unknown: while it has often been favoured in the pandemic because of lower concern for aerosolization, a disadvantage is the inability to visualize signs of tracheobronchitis and obtain biopsies. In general, more proximal samples like TA and sputum are less specific and may overestimate CAPA, although one prospective study found reasonable rates of concordance of fungal culture between TA and BAL in CAPA (16). Some centres have incorporated screening protocols for CAPA with routine measurement of serum galactomannan and/or BDG (6,17,18). While galactomannan detection in serum is likely more specific for the diagnosis of invasive disease than in bronchial fluid (depending on cut-off values), the test lacks sensitivity. Among 106 CAPA patients with serum galactomannan, 34 were positive using the recommended cut-off of 0.5 ODI; only 18 had values ≥ 1.0 ODI.

Because of the novelty of COVID-19, various research definitions for CAPA have been used in the emerging literature. Research definitions for invasive aspergillosis in other settings (such as the EORTC-MSGERC definitions (8)) are poorly-suited because biopsies – required for antemortem diagnosis of proven invasive aspergillosis – are rarely pursued in ICU patients due to risk of complications in clinically tenuous individuals, and most patients with CAPA lack underlying host factors required for classification as probable invasive aspergillosis. Consequently, most early studies used modifications of the *Asp*ICU score, adopted from studies of IAPA (4, 9). Later, in a consensus definition for IAPA, Verweij et al proposed a parallel definition for CAPA (10). White and colleagues proposed another definition (2). In December 2020, an expert panel convened by the ECMM and ISHAM proposed yet another research definition for the disease (11). Most recently, the EORTC-MSGERC Intensive Care Unit Working Group proposed definitions for invasive aspergillosis in ICU patients, including those with influenza and COVID-19 (12).

Our analysis of these definitions has identified several findings. Firstly, the incidence of CAPA may be overestimated in the literature because over a third of cases reported as CAPA did not fulfil any of these 4 standardized definitions (or were unclassifiable). For example, after pooling 21 ICU cohort studies that included patient-level data for 197 patients, the incidence of CAPA was 9.2% (as reported by authors), and this rate was considerably higher than when we reclassified these cases according to the 4 proposed standard definitions (between 1.6% and 5.2%).

Secondly, while there was overlap in the definitions, the correlation between any 2 was modest and so studies using discordant definitions may not be discussing the same patient populations. Thirdly, all definitions are hampered by lack of specificity since they all rely on clinical, radiographic, and mycological findings that may be difficult to distinguish from critically severe COVID-19.

In our study, the mortality rate of patients with CAPA was approximately 50% and was not clearly affected by antifungals. This observation remained true even when the analysis was repeated only in patients who met each of the 4 standardized research definitions for CAPA. A possible interpretation is that patients with CAPA might be diagnosed too late in their clinical course for antifungals to improve outcomes. Alternatively, for at least some patients, *Aspergillus* spp. in the airways reflects colonization and not invasive disease. A systematic review found that autopsy-proven invasive mould disease is an uncommon finding in patients dying with COVID-19, occurring in <2% of decedents reported in autopsy case series (19). Moreover, in some reported cases of probable CAPA, post-mortem examination failed to confirm the diagnosis (20). We observed a number of similarities and some contrasts with IAPA. Like with IAPA – and unlike invasive aspergillosis outside of ICU settings – most CAPA patients lacked pre-existing immunocompromising conditions. In contrast with IAPA, the timing of diagnosis of CAPA was late (occurring after a median 9 days after ICU admission), compared to after a median 2 days in IAPA (4). In addition, tracheobronchitis was rarely reported, noted in just 6% of cases in which bronchoscopy was done, in contrast to IAPA, in which it is reported to occur in one-third to one-half of affected patients (4, 10).

## Limitations

This study has some important limitations. Studies employed different surveillance methods and diagnostic tests to screen for and diagnose CAPA, and thus pooled analyses should be interpreted with caution. For example, studies that employed more aggressive surveillance – e.g. by routine bronchoscopy with fungal culture and galactomannan – would likely report higher rates of CAPA than studies in which fungal investigations were obtained only based on clinical suspicion, but conversely, may also over-represent those with *Aspergillus* spp. colonization. There was also inconsistent reporting related to clinical characteristics and diagnostics, both in patients reported to have CAPA and the rest of the ICU cohorts, limiting comparisons. For example, the use of corticosteroids and other immunomodulators were infrequently reported in CAPA and non-CAPA patients, precluding meaningful comparison. The degree to which these therapies may increase risk of CAPA is unclear (19). Finally, we did not formally assess risk of bias, and it is possible that the design of some cohort studies resulted in bias towards over- or underdiagnosing CAPA.

## Conclusions

The reported incidence of CAPA in ICU patients with COVID-19 varies greatly by study, and may be related to different surveillance protocols and inconsistent definitions which may inflate the incidence compared to standard research definitions. Although several clinical research definitions have been advanced for the diagnosis of CAPA, agreement between these is imperfect, and none clearly distinguishes which patients are likely to benefit from antifungal therapy.

## Data Availability

Not applicable; review of published data

## Contributions

RMK designed the literature search, created the figures, and wrote the first draft of the manuscript. RMK, TCD, BEK, and ISS screened and reviewed articles, and extracted and analyzed data. RMK, WIS, and ISS performed statistical analysis. ISS designed the study and was responsible for overall supervision. All authors critically reviewed and revised the manuscript and approved the final draft.

## Supplementary Data Files

Supplementary Table 1: Patient level data and reclassifications.

## References

1. World Health Organization. Coronavirus disease (COVID-19) advice for public. World Health Organization 2020 [cited 2021 May 16, 2021]. Available from: https://www.who.int/emergencies/diseases/novel-coronavirus-2019/advice-for-public

2. White PL, Dhillon R, Cordey A, Hughes H, Faggian F, Soni S, et al. A National Strategy to Diagnose Coronavirus Disease 2019–Associated Invasive Fungal Disease in the Intensive Care Unit. Clin Infect Dis. 2020 Aug 29;ciaa1298.

3. Wauters J, Baar I, Meersseman P, Meersseman W, Dams K, De Paep R, et al. Invasive pulmonary aspergillosis is a frequent complication of critically ill H1N1 patients: a retrospective study. Intensive Care Med. 2012 Nov;38(11):1761–8.

4. Schauwvlieghe AFAD, Rijnders BJA, Philips N, Verwijs R, Vanderbeke L, Van Tienen C, et al. Invasive aspergillosis in patients admitted to the intensive care unit with severe influenza: a retrospective cohort study. Lancet Respir Med. 2018 Oct;6(10):782–92.

5. Alanio A, Dellière S, Fodil S, Bretagne S, Mégarbane B. Prevalence of putative invasive pulmonary aspergillosis in critically ill patients with COVID-19. Lancet Respir Med. 2020 Jun;8(6):e48–9.

6. Brown L-AK, Ellis J, Gorton R, De S, Stone N. Surveillance for COVID-19-associated pulmonary aspergillosis. Lancet Microbe. 2020 Aug;1(4):e152.

7. Mahase E. Covid-19: WHO declares pandemic because of “alarming levels” of spread, severity, and inaction. BMJ. 2020 Mar 12;m1036.

8. Donnelly JP, Chen SC, Kauffman CA, Steinbach WJ, Baddley JW, Verweij PE, et al. Revision and Update of the Consensus Definitions of Invasive Fungal Disease From the European Organization for Research and Treatment of Cancer and the Mycoses Study Group Education and Research Consortium. Clin Infect Dis. 2020 Sep 12;71(6):1367–76.

9. Blot SI, Taccone FS, Van den Abeele A-M, Bulpa P, Meersseman W, Brusselaers N, et al. A Clinical Algorithm to Diagnose Invasive Pulmonary Aspergillosis in Critically Ill Patients. Am J Respir Crit Care Med. 2012 Jul;186(1):56–64.

10. Verweij PE, Gangneux J-P, Bassetti M, Brüggemann RJM, Cornely OA, Koehler P, et al. Diagnosing COVID-19-associated pulmonary aspergillosis. Lancet Microbe. 2020 Jun;1(2):e53– 5.

11. Koehler P, Bassetti M, Chakrabarti A, Chen SCA, Colombo AL, Hoenigl M, et al. Defining and managing COVID-19-associated pulmonary aspergillosis: the 2020 ECMM/ISHAM consensus criteria for research and clinical guidance. Lancet Infect Dis. 2020 Dec;S1473309920308471.

12. Bassetti M, Azoulay E, Kullberg B-J, Ruhnke M, Shoham S, Vazquez J, et al. EORTC/MSGERC Definitions of Invasive Fungal Diseases: Summary of Activities of the Intensive Care Unit Working Group. Clin Infect Dis. 2021 Mar 12;72(Supplement_2):S121–7.

13. Moola S, Munn Z, Tufanaru C, Aromataris E, Sears K, Sfetc R, et al. Chapter 7: Systematic Reviews of Etiology and Risk. In: Aromataris E, Munn Z, editors. JBI Manual for Evidence Synthesis [Internet]. JBI; 2020 [cited 2021 May 17]. Available from: https://wiki.jbi.global/display/MANUAL/Chapter+7%3A+Systematic+reviews+of+etiology+and+risk

14. Bartoletti M, Pascale R, Cricca M, Rinaldi M, Maccaro A, Fornaro G, et al. Epidemiology of invasive pulmonary aspergillosis among COVID-19 intubated patients: a prospective study. :31.

15. Wang J, Yang Q, Zhang P, Sheng J, Zhou J, Qu T. Clinical characteristics of invasive pulmonary aspergillosis in patients with COVID-19 in Zhejiang, China: a retrospective case series. Crit Care. 2020 Dec;24(1):299.

16. Garcia-Vidal C, Sanjuan G, Moreno-García E, Puerta-Alcalde P, Garcia-Pouton N, Chumbita M, et al. Incidence of co-infections and superinfections in hospitalized patients with COVID-19: a retrospective cohort study. Clin Microbiol Infect. 2021 Jan;27(1):83–8.

17. Helleberg M, Steensen M, Arendrup MC. Invasive aspergillosis in patients with severe COVID-19 pneumonia. Clin Microbiol Infect. 2021 Jan;27(1):147–8.

18. van Arkel ALE, Rijpstra TA, Belderbos HNA, van Wijngaarden P, Verweij PE, Bentvelsen RG. COVID-19-associated Pulmonary Aspergillosis. Am J Respir Crit Care Med. 2020 Jul 1;202(1):132–5.

19. Kula BE, Clancy CJ, Nguyen MH, Schwartz IS. Invasive Mould Disease in Fatal COVID-19: A Systematic Review of Autopsies; 2021 Jan [cited 2021 May 16]. Available from: http://medrxiv.org/lookup/doi/10.1101/2021.01.13.21249761

20. Grootveld R, Paassen J, Boer MGJ, Claas ECJ, Kuijper EJ, Beek MT, et al. Systematic screening for COVID□19 associated invasive aspergillosis in ICU patients by culture and PCR on tracheal aspirate. Mycoses. 2021 Jun;64(6):641–50.

21. Dupont D, Menotti J, Turc J, Miossec C, Wallet F, Richard J-C, et al. Pulmonary aspergillosis in critically ill patients with Coronavirus Disease 2019 (COVID-19). Med Mycol. 2021 Jan 4;59(1):110–4.

22. Lamoth F, Glampedakis E, Boillat-Blanco N, Oddo M, Pagani J-L. Incidence of invasive pulmonary aspergillosis among critically ill COVID-19 patients. Clin Microbiol Infect. 2020 Dec;26(12):1706–8.

23. Nasir N, Farooqi J, Mahmood SF, Jabeen K. COVID□19□associated pulmonary aspergillosis (CAPA) in patients admitted with severe COVID□19 pneumonia: An observational study from Pakistan. Mycoses. 2020 Aug;63(8):766–70.

24. Rutsaert L, Steinfort N, Van Hunsel T, Bomans P, Naesens R, Mertes H, et al. COVID-19-associated invasive pulmonary aspergillosis. Ann Intensive Care. 2020 Dec;10(1):71.

25. Van Biesen S, Kwa D, Bosman RJ, Juffermans NP. Detection of Invasive Pulmonary Aspergillosis in COVID-19 with Nondirected BAL. Am J Respir Crit Care Med. 2020 Oct 15;202(8):1171–3.

26. Lahmer T, Rasch S, Spinner C, Geisler F, Schmid RM, Huber W. Invasive pulmonary aspergillosis in severe coronavirus disease 2019 pneumonia. Clin Microbiol Infect. 2020 Oct;26(10):1428–9.

27. Ichai P, Saliba F, Baune P, Daoud A, Coilly A, Samuel D. Impact of negative air pressure in ICU rooms on the risk of pulmonary aspergillosis in COVID-19 patients. Crit Care. 2020 Dec;24(1):538.

28. Gangneux J-P, Reizine F, Guegan H, Pinceaux K, Le Balch P, Prat E, et al. Is the COVID-19 Pandemic a Good Time to Include Aspergillus Molecular Detection to Categorize Aspergillosis in ICU Patients? A Monocentric Experience. J Fungi. 2020 Jul 10;6(3):105.

29. Sarrazyn C, Dhaese S, Demey B, Vandecasteele S, Reynders M, Van Praet JT. Incidence, risk factors, timing and outcome of influenza versus Covid-19 associated putative invasive aspergillosis. Infect Control Hosp Epidemiol. 2020 Sep 9;1–7.

30. Agrifoglio A, Cachafeiro L, Figueira JC, Añón JM, García de Lorenzo A. Critically ill patients with COVID-19 and candidaemia: We must keep this in mind. J Mycol Médicale. 2020 Dec;30(4):101012.

31. Ripa M, Galli L, Poli A, Oltolini C, Spagnuolo V, Mastrangelo A, et al. Secondary infections in patients hospitalized with COVID-19: incidence and predictive factors. Clin Microbiol Infect. 2021 Mar;27(3):451–7.

32. Versyck M, Zarrougui W, Lambiotte F, Elbeki N, Saint-Leger P. Invasive pulmonary aspergillosis in COVID-19 critically ill patients: Results of a French monocentric cohort. J Med Mycol. 2021 Jun;31(2):101122.

33. Rothe K, Feihl S, Schneider J, Wallnöfer F, Wurst M, Lukas M, et al. Rates of bacterial co-infections and antimicrobial use in COVID-19 patients: a retrospective cohort study in light of antibiotic stewardship. Eur J Clin Microbiol Infect Dis. 2021 Apr;40(4):859–69.

34. Maes M, Higginson E, Pereira-Dias J, Curran MD, Parmar S, Khokhar F, et al. Ventilator-associated pneumonia in critically ill patients with COVID-19. Crit Care. 2021 Dec;25(1):25.

35. Dellière S, Dudoignon E, Fodil S, Voicu S, Collet M, Oillic P-A, et al. Risk factors associated with COVID-19-associated pulmonary aspergillosis in ICU patients: a French multicentric retrospective cohort. Clin Microbiol Infect. 2021 May;27(5):790.e1-790.e5.

36. Permpalung N, Chiang TP-Y, Massie AB, Zhang SX, Avery RK, Nematollahi S, et al. COVID-19 Associated Pulmonary Aspergillosis in Mechanically Ventilated Patients. Clin Infect Dis. 2021 Mar 9;ciab223.

37. Segrelles□Calvo G, Araújo GRS, Llopis□Pastor E, Carrillo J, Hernández□Hernández M, Rey L, et al. Prevalence of opportunistic invasive aspergillosis in COVID_19 patients with severe pneumonia. Mycoses. 2021 Feb;64(2):144–51.

38. Razazi K, Arrestier R, Haudebourg AF, Benelli B, Carteaux G, Decousser J, et al. Risks of ventilator-associated pneumonia and invasive pulmonary aspergillosis in patients with viral acute respiratory distress syndrome related or not to Coronavirus 19 disease. Crit Care. 2020 Dec;24(1):699.

39. Roman□Montes CM, Martinez□Gamboa A, Diaz□Lomelí P, Cervantes□Sanchez A, Rangel□Cord A, Sifuentes□Osorni J, et al. Accuracy of galactomannan testing on tracheal aspirates in COVID□19□associated pulmonary aspergillosis. Mycoses. 2021 Apr;64(4):364–71.

40. Machado M, Valerio M, Álvarez□Uría A, Olmedo M, Veintimilla C, Padilla B, et al. Invasive pulmonary aspergillosis in the COVID□19 era: An expected new entity. Mycoses. 2021 Feb;64(2):132–43.

41. Meijer EFJ, Dofferhoff ASM, Hoiting O, Meis JF. COVID□19–associated pulmonary aspergillosis: a prospective single□center dual case series. Mycoses. 2021 Apr;64(4):457–64.

42. Søgaard KK, Baettig V, Osthoff M, Marsch S, Leuzinger K, Schweitzer M, et al. Community-acquired and hospital-acquired respiratory tract infection and bloodstream infection in patients hospitalized with COVID-19 pneumonia. J Intensive Care. 2021 Dec;9(1):10.

43. Borman AM, Palmer MD, Fraser M, Patterson Z, Mann C, Oliver D, et al. COVID-19-Associated Invasive Aspergillosis: Data from the UK National Mycology Reference Laboratory. Hanson KE, editor. J Clin Microbiol. 2020 Dec 17;59(1):JCM.02136-20, e02136-20.

44. Fekkar A, Lampros A, Mayaux J, Poignon C, Demeret S, Constantin J-M, et al. Occurrence of Invasive Pulmonary Fungal Infections in Patients with Severe COVID-19 Admitted to the ICU. Am J Respir Crit Care Med. 2021 Feb 1;203(3):307–17.

45. Boyd S, Martin-Loeches I. Rates of Aspergillus Co-infection in COVID patients in ICU not as high as previously reported. Clin Infect Dis. 2021 Jan 8;ciab008.

46. Marr KA, Platt A, Tornheim JA, Zhang SX, Datta K, Cardozo C, et al. Aspergillosis Complicating Severe Coronavirus Disease. Emerg Infect Dis. 2021 Jan;27(1):18–25.

47. Mitaka H, Perlman DC, Javaid W, Salomon N. Putative invasive pulmonary aspergillosis in critically ill patients with COVID□19: An observational study from New York City. Mycoses. 2020 Dec;63(12):1368–72.

48. Benedetti MF, Alava KH, Sagardia J, Cadena RC, Laplume D, Capece P, et al. COVID-19 associated pulmonary aspergillosis in ICU patients: Report of five cases from Argentina. Med Mycol Case Rep. 2021 Mar;31:24–8.

